# COVID-19 pandemic re-shaped the global dispersal of seasonal influenza viruses

**DOI:** 10.1101/2023.12.20.23300299

**Authors:** Zhiyuan Chen, Joseph L.-H. Tsui, Bernardo Gutierrez, Simon Busch Moreno, Louis du Plessis, Xiaowei Deng, Jun Cai, Sumali Bajaj, Marc A. Suchard, Oliver G. Pybus, Philippe Lemey, Moritz U. G. Kraemer, Hongjie Yu

## Abstract

Understanding how the global dispersal patterns of seasonal influenza viruses were perturbed during and after the COVID-19 pandemic is needed to inform influenza intervention and vaccination strategies in the post-pandemic period. Although global human mobility has been identified as a key driver of influenza dispersal^1^, alongside climatic and evolutionary factors^2,3^, the impact of international travel restrictions on global influenza transmission and recovery remains unknown. Here we combine molecular, epidemiological, climatic, and international travel data within a phylodynamic framework to show that, despite human mobility remaining the principal driver of global influenza virus dissemination, the pandemic’s onset led to a shift in the international population structure and migration network of seasonal influenza lineages. We find that South Asia and Africa played important roles as exporters and phylogenetic trunk locations of influenza in 2020 and 2021, and we highlight the association between population movement, antigenic drift and persistence during the intensive non-pharmaceutical interventions (NPIs) phase. The influenza B/Yamagata lineage disappeared in a context of reduced relative genetic diversity, moderate lineage turnover, and lower positive selection pressure. Our results demonstrate that mobility perturbations reshaped the global dispersal dynamics of influenza viruses, with potential implications for vaccine design and genomic surveillance programmes. As the risk of future pandemics persists, our study provides an opportunity to assess the impact of NPIs during the pandemic on respiratory infectious diseases beyond the interplay between SARS-CoV-2 and influenza viruses.

## Main

Seasonal influenza epidemics place substantial burdens on healthcare systems and cause >5 million hospitalizations among adults every year^4^. The global spread of recurring seasonal influenza is strongly associated with global air travel^1,5-7^ as well as with the periodic antigenic evolution of influenza viruses that permit immune escape from vaccine- and/or infection-induced immunity^8^.

The implementation of non-pharmaceutical interventions (NPIs) in 2020 in response to the COVID-19 pandemic affected the evolution and circulation of seasonal influenza viruses^9^ and other respiratory pathogens^10^. Following the introduction of NPIs, a rapid decline in influenza transmission was observed worldwide, leading to changes in accumulation of population immunity and substantial genetic bottlenecks constraining virus diversity^8^. Influenza transmission and dispersal resurged in late 2021 after the relaxation of NPIs, but one influenza B virus lineage, B/Yamagata, has been reported only rarely since March 2020^11^. This led the World Health Organization (WHO) to recommend using trivalent vaccines that exclude the B/Yamagata strain in the 2024 southern hemisphere influenza season^12^.

The current paradigm for influenza vaccine development emphasises timely evaluation of the antigenic and genetic characteristics of circulating strains^13^, especially those sampled from previously identified source populations (including subtropical and tropical East Asia, Southeast Asia, and occasionally India for A/H3N2)^14,15^. We hypothesised that the NPIs implemented during the COVID-19 pandemic reshaped the spatial dissemination and evolutionary patterns of seasonal influenza in a geographically-heterogeneous manner.

By combining epidemiological, genetic, and international travel data together in a phylodynamic framework, we estimate the spatio-temporal population structure, persistence patterns, and evolutionary diversity of seasonal influenza before, during, and after the NPIs-intensive phase of the COVID-19 pandemic.

## Results

### Decline in global influenza transmission

Using the COVID-19 stringency index and monthly air passenger volumes over time (**Extended Data Fig. 1**), we defined three “periods” with distinct patterns of global human mobility: period 1 is prior to population-level behavioural changes in response to the COVID-19 pandemic (1 February 2018 - 31 January 2020), period 2 is characterised by high-stringency interventions (referred to as NPIs-intensive phase) that aimed to limit population mixing between 1 February 2020 and 31 July 2021, and period 3 covers the time when international travel mostly had recovered (1 August 2021 to 31 July 2023).

We obtained influenza virological surveillance data from the Global Influenza Surveillance and Response System (GISRS)^16^, which collates data from specimens from patients with influenza-like illness that have been tested for influenza viruses at National Influenza Centres, national influenza reference laboratories, and other non-sentinel systems^17^. The antigenic and genetic characterisation of influenza viruses collected via this network form the basis of WHO annual recommendations for the composition of influenza vaccines. The number of specimens processed for influenza testing remained stable during the NPIs-intensive phase compared to previous influenza seasons, and doubled in 2022 and 2023 (**Fig. 1a**). This rise is possibly due to an increase in the global capacity for viral surveillance established during the pandemic, and a re-focus on influenza sentinel surveillance as COVID-19 cases subsided ^18^. The proportion of laboratory confirmed influenza cases that were subsequently sequenced during the NPIs-intensive phase of the pandemic was generally above 10%; this proportion was lower in 2019 and 2022, 2023 (**Fig. 1b**). Even though surveillance intensity for seasonal influenza was maintained during the pandemic, we cannot rule out possible biases inherent to influenza databases of virological and genomic surveillance^16,19^.

**Fig 1.**
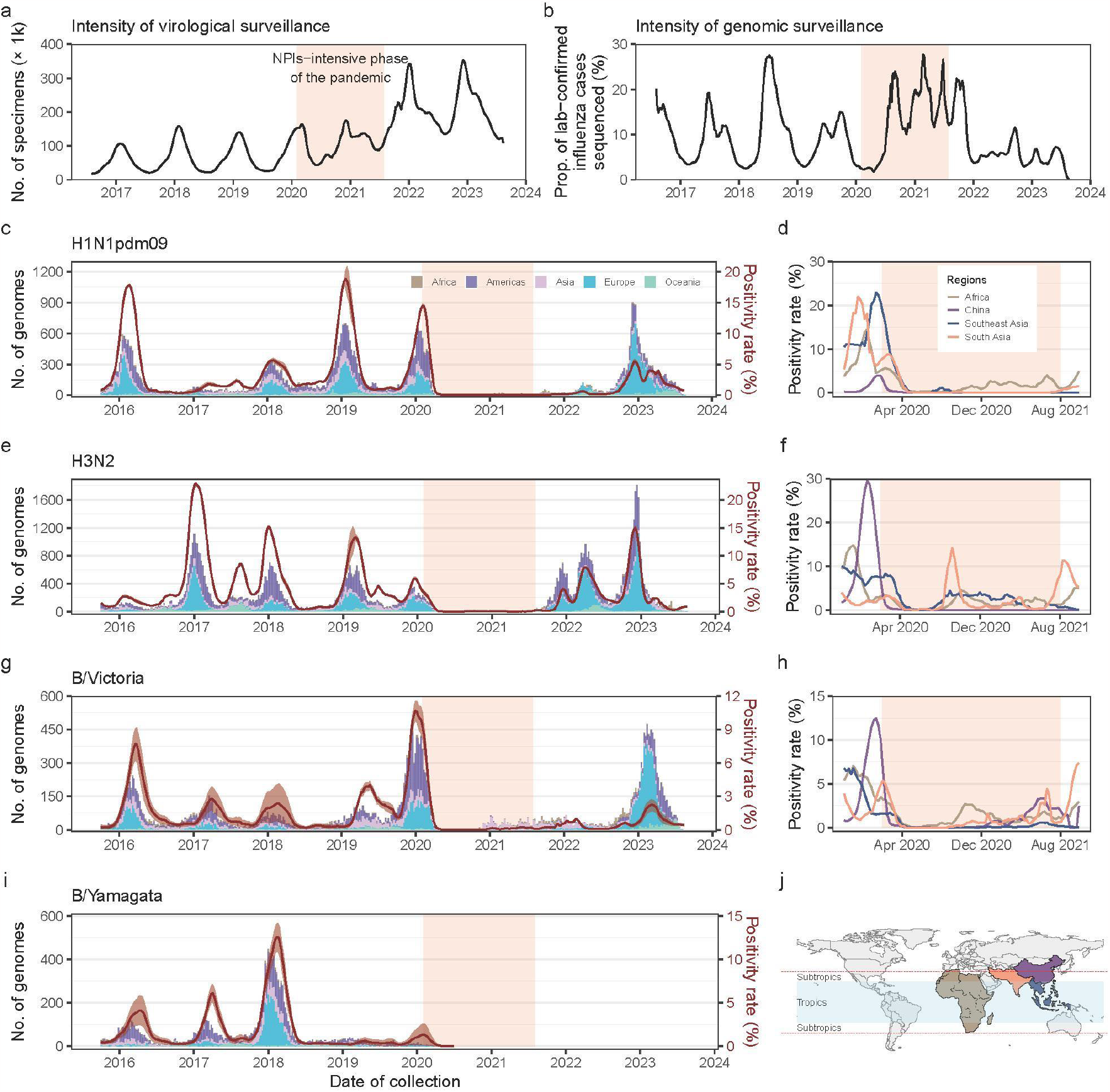
Surveillance intensity, distribution of genetic sequences, and positivity rates of seasonal influenza viruses. **a**, Intensity of virological surveillance of influenza indicated by the rolling numbers of specimens processed globally for the test of influenza virus. **b**, Intensity of genomic surveillance of influenza indicated by the rolling percentages of reported influenza cases sequenced at high quality. **c-i**, The weekly count of high-quality genetic sequences (HA segment, stratified by continent) and global positivity rates among tested specimens for H1N1pdm09 (**c**), H3N2 (**e**), B/Victoria (**g**) and B/Yamagata (**i**). The positivity rates of H1N1pdm09 (**d**), H3N2 (**f**), B/Victoria (**h**) are also presented separately for regions that experienced influenza waves during the NPIs-intensive phase of pandemic (Africa, China, Southeast Asia, and South Asia). **j**, The geographic location of Africa, China, Southeast Asia, and South Asia in reference to tropical and subtropical latitudinal limits. Positivity rates are presented as central-aligned rolling averages (5-week window), and the 95% intervals indicate uncertainty in inferring the specific subtypes or lineages using a Bayesian framework. The light orange shadow area represents the NPIs-intensive phase of the COVID-19 pandemic, defined as spanning from 1 February 2020 to 31 July 2021.

The fraction of specimens that tested positive for influenza viruses remained low during the NPIs-intensive period (< 0.10% for H1N1pdm09, <0.25% for H3N2, and <0.50% for B/Victoria in each week from May 2020 to July 2021), compared to the peak positivity rates of H1N1pdm09 (14.5%), H3N2 (6.0%), and B/Victoria (10.7%) during the 2019-2020 northern hemisphere winter (**Fig. 1c, 1e, 1g**). To account for variation in the number of specimens across the three periods, we also calculated the ratio of positive influenza cases to the total number of specimens processed in each period. These data also indicate low levels of influenza transmission after the emergence of SARS-CoV-2 (**Extended Data Fig. 2**). However, several small-scale influenza outbreaks occurred in Asia (mainly in South Asia, Southeast Asia, and China) and Africa in period 2 (**Fig. 1d, 1f, 1h, 1j**), both in areas with high and low COVID-19 burden and varying levels of population mixing. A H3N2 wave occurred during the 2021-2022 northern hemisphere winter season, with a double peak before and after the winter holidays, coinciding with large waves of SARS-CoV-2 Omicron BA.1 and BA.2 infection^20^. This H3N2 wave was followed by a H1N1pdm09 wave in the first half of 2022 and a B/Victoria wave, although low-level local circulation of B/Victoria continued across Asia (mainly China and South Asia) and Africa during the NPIs-intensive phase (**Fig. 1c-h**). A few B/Yamagata cases were sporadically reported after the onset of pandemic, however these might originate from vaccine-derived cases or potential data errors^11,21^. Substantial transmission of H1N1pdm09, H3N2, and B/Victoria occurred in the 2022-2023 northern hemisphere winter season, during which weekly positivity rates peaked at 15.0% for H3N2 (**Fig. 1c-h**).

### Air passenger traffic as a predictor of influenza spread

The spatial spread of influenza virus has been shown previously to be associated with climatic factors (primarily humidity)^22^ and global human mobility^1^. Disruptions to air travel, like those during September 2001 in the United States, have been shown to impact the timing of subsequent influenza season^23^. To infer period-specific predictors and to account for potential variation in rates of viral dispersal across the three periods defined above, we constructed a 3-period Bayesian phylogeographic generalised linear model (GLM) framework and applied it to clades of influenza viruses circulating from November 2016 (September 2013 for B/Yamagata) to July 2023 (see **Methods**). To account for potential biases in genetic sampling, we adopted three sub-sampling strategies for high-quality genetic sequences (hemagglutinin, HA, segment) (**Supplementary Fig 1**): (i) even sub-sampling, (ii) sub-sampling while accounting for temporal variation, and (iii) sub-sampling while accounting for both temporal and spatial variation (see **Methods**). These schemes were used to generate sets of ∽2,000 sequences for each influenza subtype/lineage, with collection dates up to 31 July 2023 (**Extended Data Fig. 3**) and sampled from 12 geographic regions (Africa, North America, South America, Europe, Russia, North China, South China, Japan/Korea, South Asia, West Asia, Southeast Asia, and Oceania; **Supplementary Fig 2**). Covariates related to demographic, meteorological, air passenger traffic, geographic, and sampling factors were included in the GLM model to explore their period-specific associations with virus lineage movements (**Supplementary Fig 3-5, Supplementary Table 1-3**).

We observed heterogeneous patterns in the reduction of air passenger traffic between each pair of regions during the pandemic (**Fig. 2a-f, Supplementary Fig 6**), although the structure of the airline network did not change fundamentally (Mantel statistic R between period 1 and period 2: 0.941, *p* < 0.01; between period 1 and period 3: 0.917, *p* < 0.01). During the second period, the most severe reduction in bidirectional air traffic occurred between China and other regions. During this time air traffic between North China and Russia, North America, Southeast Asia, South America, Oceania, and Europe all dropped to <7.5% of pre-pandemic levels (**Fig. 2e**). Following the relaxation of NPIs (period 3), the rate of recovery of air traffic volumes also varied across regions, with slower recoveries in Asia (e.g., Southeast Asia, China, Japan/Korea) and Oceania (**Fig. 2d**). Average monthly air passenger traffic during the third period had not recovered to >10.0% of pre-pandemic levels on routes between North China and North America, and between Japan/Korea and Russia (**Fig. 2f**).

**Fig 2.**
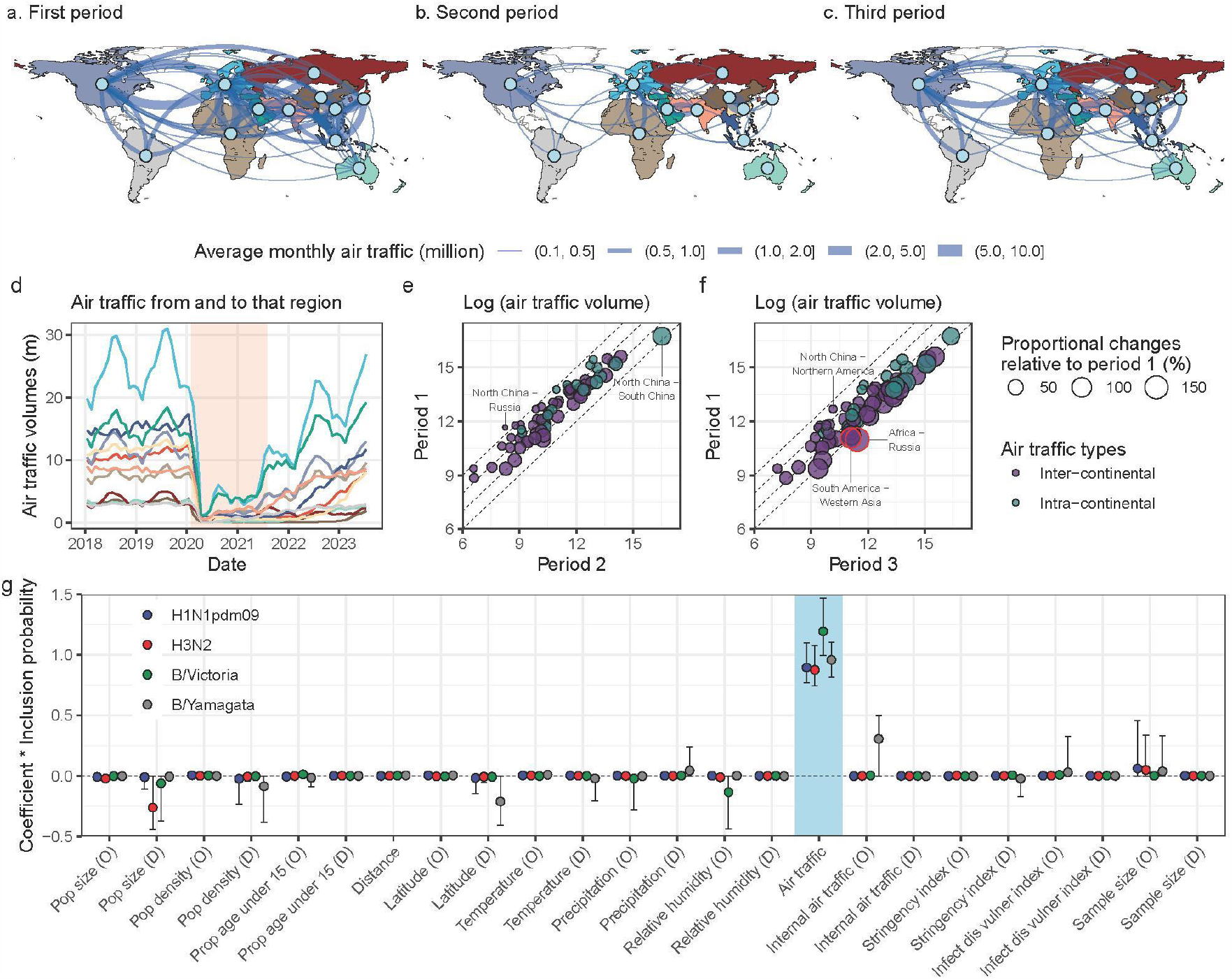
Predictors of global movements of seasonal influenza virus using a 3-period phylogeographic GLM model. **a-c**, Average monthly air passenger traffic network between 12 geographic regions across three periods. Here, routes with average monthly air traffic passengers of more than 100,000 are only presented for a clear visualisation. **d**, Region-specific monthly air traffic from and to that region over time. The air traffic between south China and north China was not included in this calculation. Colours correspond to the coloured background of the global map in panel **a-c. e-f**, Using first period as baseline, comparison of average monthly bidirectional passenger air traffic volume in each pair of regions with that in the second and third period, respectively. Bidirectional air traffic routes among 12 regions were presented as points coloured by the intercontinental or intracontinental type. Routes with more than 100% recovery are marked in a red frame. **g**, Posterior summaries of the product of the log predictor inclusion probability pooled across the three periods and constant-through-time predictor coefficient in GLM model for H1N1pdm09, H3N2, and B/Victoria influenza virus. Analyses for B/Yamagata virus were performed under a time-homogeneous (1-period) GLM model since few genetic data available after March 2020. Points and ranges represent the posterior mean and 95% highest posterior density (HPD) intervals, respectively. Location-specific predictors were included for both origin (O) and destination (D) locations in the pairwise transition rates.

Despite changes in travel volumes, air passenger traffic among regions remained a significant predictor of the global spread of influenza across all periods (**Fig. 2g;** results consistent among three sub-sampling schemes; **Extended Data Fig. 4**). As sample size had a lower predictive power under the “even sub-sampling” scheme, we chose that sampling scheme for the main analysis, similar to a previous study^15^. Climatic and demographic factors were not identified as significant predictors for influenza spread, which we hypothesise might be a consequence of the high level of spatial aggregation of the predictor variables.

### Reshaping global influenza circulation dynamics

Changes in human behaviours and air passenger traffic during the pandemic may have perturbed the local and global circulation dynamics of seasonal influenza. To investigate this, we used time-variable air traffic volumes as a sole predictor of among-location lineage transition rates and extended the phylogeographic model to also include overall air traffic as a predictor for the overall transition rates (see **Methods, Supplementary Table 4**). We first estimated trends through time in the number of lineage movement events (**Fig. 3a**). Second, we investigated how the among-location migration network changed across all periods (**Fig. 3b-e, Extended Data Fig. 5-6**) and computed changes in net lineage export events (export event count minus import event count), per region per period (**Extended Data Fig. 7**). Third, we inferred the time-varying location of the trunk phylogenetic branch based on our phylogeographic analysis (**Fig. 4**). As in previous studies, this branch represents the lineage(s) that successfully persists from one epidemic season to the next, under the defined sampling scheme^1^.

**Fig 3.**
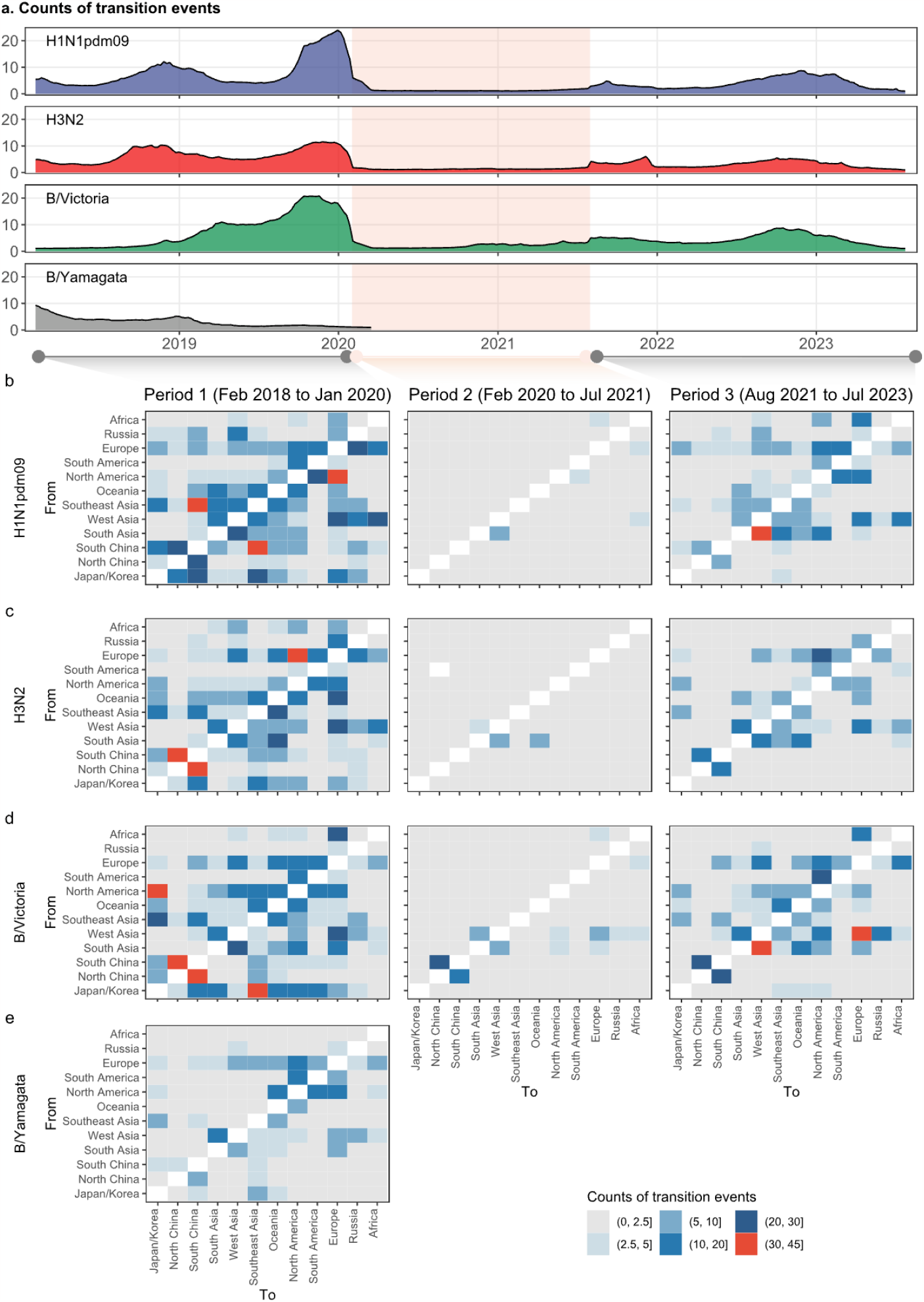
The migration dynamic of seasonal influenza virus across the three periods. **a**, The weekly counts of Markov jumps (location transition events) over time for four influenza subtypes/lineages. **b-e**, Estimates of location transition events between each pair of the geographic regions in three periods under the even sub-sampling scheme. Analyses are based on the posterior summaries of the Markov jumps under a time-inhomogeneous (3-period) GLM model with only air traffic data as the predictor of overall and relative transition rates, except for B/Yamagata under a time-homogeneous (1-period) GLM model.

**Fig 4.**
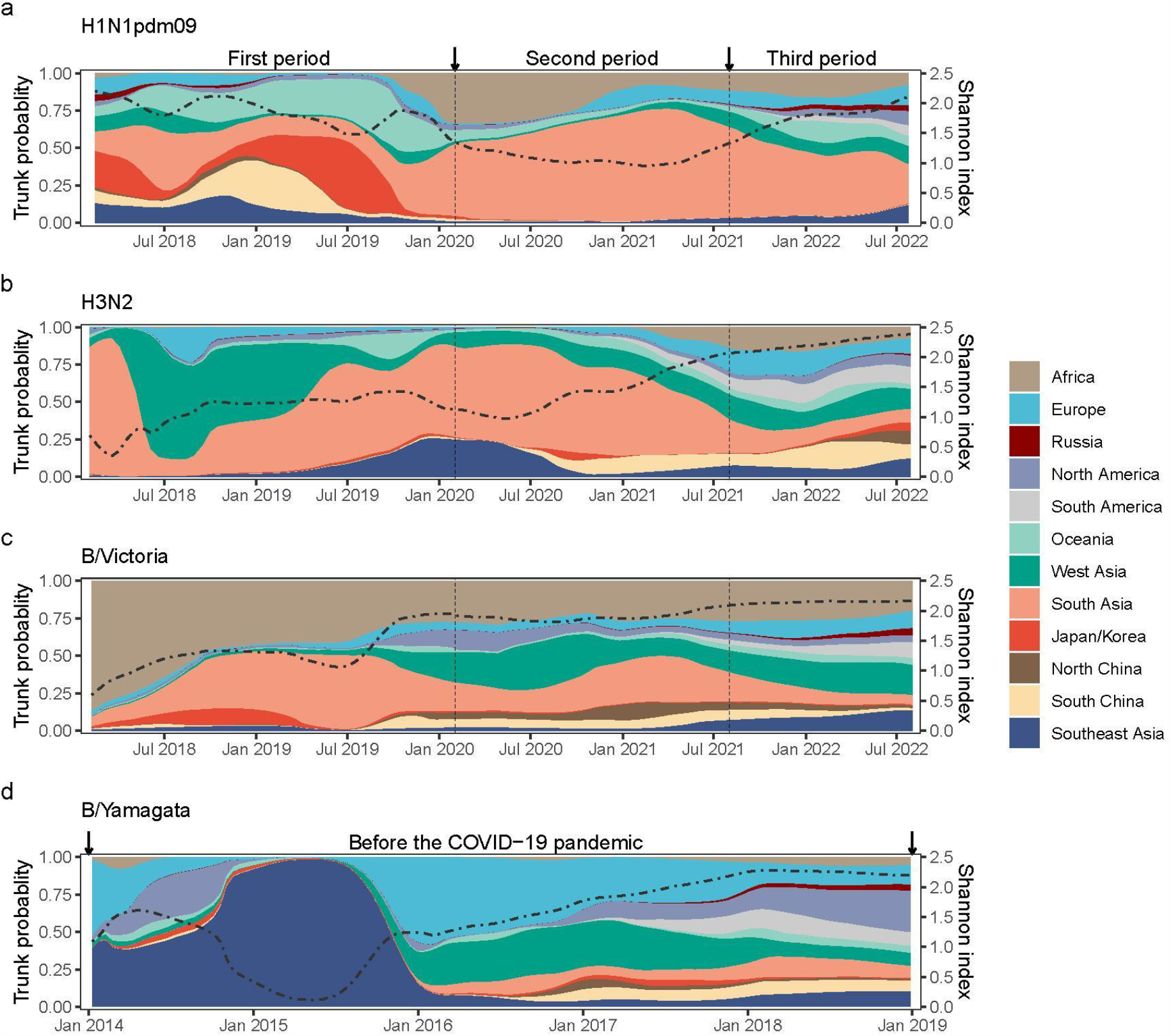
Inferred trunk location of phylogenetic trees during the three periods. Trunk probability refers to the lineage that successfully persists from one epidemic season to the next under our sampling framework. The trunk probabilities through time are averaged over two subsampling strategies (even sub-sampling, sub-sampling accounting for temporal variation; see Methods). Variation in trunk lineage diversity is represented by the Shannon index (dashed black line, second y axis; this value increases as both the number of trunk lineages and the evenness of their frequencies increase).

The H1N1pdm09, H3N2, and B/Victoria lineages all exhibited seasonal fluctuations in the number of among-location movements, while the B/Yamagata lineage showed fewer and declining events before the pandemic (**Fig. 3a, Extended Data Fig. 5-6**). During period 1 we identified a higher number of H1N1pdm09 lineage movements between Southeast Asia and South China (**Fig. 3b**), as well as a higher number of H3N2 and B/Victoria transitions between northern and southern China (**Fig. 3c-d**). Conversely, few B/Yamagata movements originated from Asia (**Fig. 3e**). In period 2 virus movements decreased substantially, although some B/Victoria movements between northern and southern China were still observed (**Fig. 3d**). This could be related to (i) B/Victoria’s greater local persistence between seasons^15^, (ii) its acquisition of adaptive amino acid mutations during intensive NPIs^24^, and (iii) higher relative human mobility between these regions due to low local COVID-19 prevalence (at a time when prevalence was higher in other world regions). During period 3, there was some re-emergence of lineage movement, which varied across regions and virus subtypes/lineages (**Fig. 3, Extended Data Fig. 5-6**). Generally, a shift to higher virus movements from South Asia to West Asia occurred, especially for H1N1pdm09 and B/Victoria (**Fig. 3b-d**). Unlike virus lineage movements from Asia, which occurred at high rates after the peak of pandemic restrictions in early August 2021 (**Extended Data Fig. 5a-c**), lineage movements from North America were limited until late 2022; this is most evident for influenza A viruses, which showed limited spread from North America to Europe during the summer of 2021 (**Extended Data Fig. 5-6**). Lineage movements among regions returned to pre-pandemic patterns by mid-2022 (**Extended Data Fig. 5**), although the absolute number of observed movements was lower than before the pandemic (**Fig 3a, Extended Data Fig. 5**).

Period- and region-specific lineage exports were heterogeneous among locations and time periods. Based on our phylogeographic model, we inferred that Asia was the main net exporter of H1N1pdm09 and B/Victoria lineages in period 1. However, within Asia there was a shift towards lineage net export from South Asia after the NPI-intensive phase of the pandemic (**Extended Data Fig. 7a, 7c**). South Asia became the sole source of observed net lineage exports of H3N2 in period 2; Europe was identified as a notable net exporter in period 3, and meanwhile more importations occurred in Southeast Asia than exportations in that period (**Extended Data Fig. 7b**). This shift may be the consequence of the timing and intensity of reopening of European air travel compared to that in Asia.

For the two years before the start of the COVID-19 pandemic, we infer that H1N1pdm09 trunk lineages were most likely located in East-Southeast Asia and South Asia. Although the posterior probability of the trunk lineage being in Africa grew after 2019, the posterior probability for the location of the H1N1pdm09 trunk lineage was greatest for South Asia in the second and third periods (**Fig. 4a**), consistent with the net export results described above (**Extended Data Fig. 7a**). For H3N2, we also inferred Asia as the most likely trunk location during period 1. During period 2, South Asia was the most likely H3N2 trunk location, with a reduced posterior probability for Southeast Asia (**Fig. 4b**). South Asia, West Asia and Africa together contributed >60% of the posterior probability for the B/Victoria trunk location (**Fig. 4c**). The B/Yamagata trunk lineage in the pre-pandemic period was most likely in Southeast Asia in 2014-2015 and Europe in 2016 (**Fig. 4d**).

### Heterogeneous persistence patterns and potential drivers

Measures of local persistence of influenza viruses between seasons can help predict the global distribution of circulating strains and are used to inform vaccine compositions^14^. To investigate drivers of persistence during the COVID-19 pandemic, we analyse and contrast two measures of persistence across all three periods^15,25^: i) tip-associated persistence, which represents the time that terminal branches spend in location of the sampled tip, when moving backwards through the tree, from the sampled tip towards the root (**Methods**), ii) lineage-associated persistence, which represents the time that each lineage - which is present at a given evaluation time - spends in its current location, when moving backwards through the tree, towards the root (**Methods**).

Tip-associated persistence of influenza viruses during the first period was generally <6 months, although greater persistence was observed for B/Yamagata (**Extended Data Fig. 8**). In period 2, influenza viruses exhibited longer persistence, likely due to NPIs reducing long-distance travel and lineage movement (**Fig. 3**) and subsequent strain replacement. For example, a high tip-associated persistence was observed for H3N2 viruses circulating in Africa (1.79 years, interquartile range: 1.42-2.14), South Asia (1.40, 1.25-1.70), and Southeast Asia (0.85, 0.64-1.21, **Extended Data Fig. 8b**). Lineage-associated persistence shows similar trends for H3N2, which was the only subtype with sufficient data to estimate temporal trends across regions during the second period (**Extended Data Fig. 9a, Supplementary Fig. 7**).

Using a Bayesian hierarchical regression model (**Methods**), we found that air traffic reduction and antigenic drift (measured using amino acid-based epitope distances^26^) were positively associated with lineage-associated persistence of H3N2 viruses. During the second period, we found that the association between antigenic drift and persistence increased in Africa (z-score: 0.74, 90% highest density intervals (HDIs): 0.55-0.91), and that as persistence increased, air traffic decreased (z-score: 1.05, 0.87-1.23) (**Extended Data Fig. 9b**). Similar associations between persistence and air traffic reduction were observed in South Asia (**Extended Data Fig. 9b**).

During the third period, as population mixing increased and international virus lineage movements resurged, global circulation was re-established resulting in a drop in persistence. However, high persistence of influenza viruses was still observed in some regions (e.g., H1N1pdm09 in Africa and South Asia; H3N2 in China and South Asia; B/Victoria in Africa and China, **Extended Data Fig. 8a-c**), potentially because of slower recovery of air passenger traffic (**Fig. 2d**). As transmission of B/Victoria resumed in the third period, tip-associated persistence increased in some regions, confirming its relative ability in persisting locally^15^ (**Extended Data Fig. 8c**).

Measures of tip- and lineage-associated persistence used here are based on phylogenetic analyses and cannot be interpreted as equivalent to the persistence of transmission chains across seasons, due to the impact of sampling biases of phylogenetic topology and uncertainty in phylogeographic inferences^25,27^. New models that integrate phylogenetic analyses with mathematical models of transmission of influenza could help bridge this gap.

### Temporal patterns of genetic diversity

Although the global dispersal of influenza A virus subtypes and B/Victoria lineage shifted during the COVID-19 pandemic, the circulation of B/Yamagata lineages likely ceased from early 2020 onwards (**Fig. 5a**). Given that the shared evolutionary history of some influenza B virus segments across the B/Victoria and B/Yamagata lineages, we focussed on the HA segment, one of three segments experiencing distinct evolution in B lineages^28^.

**Fig 5.**
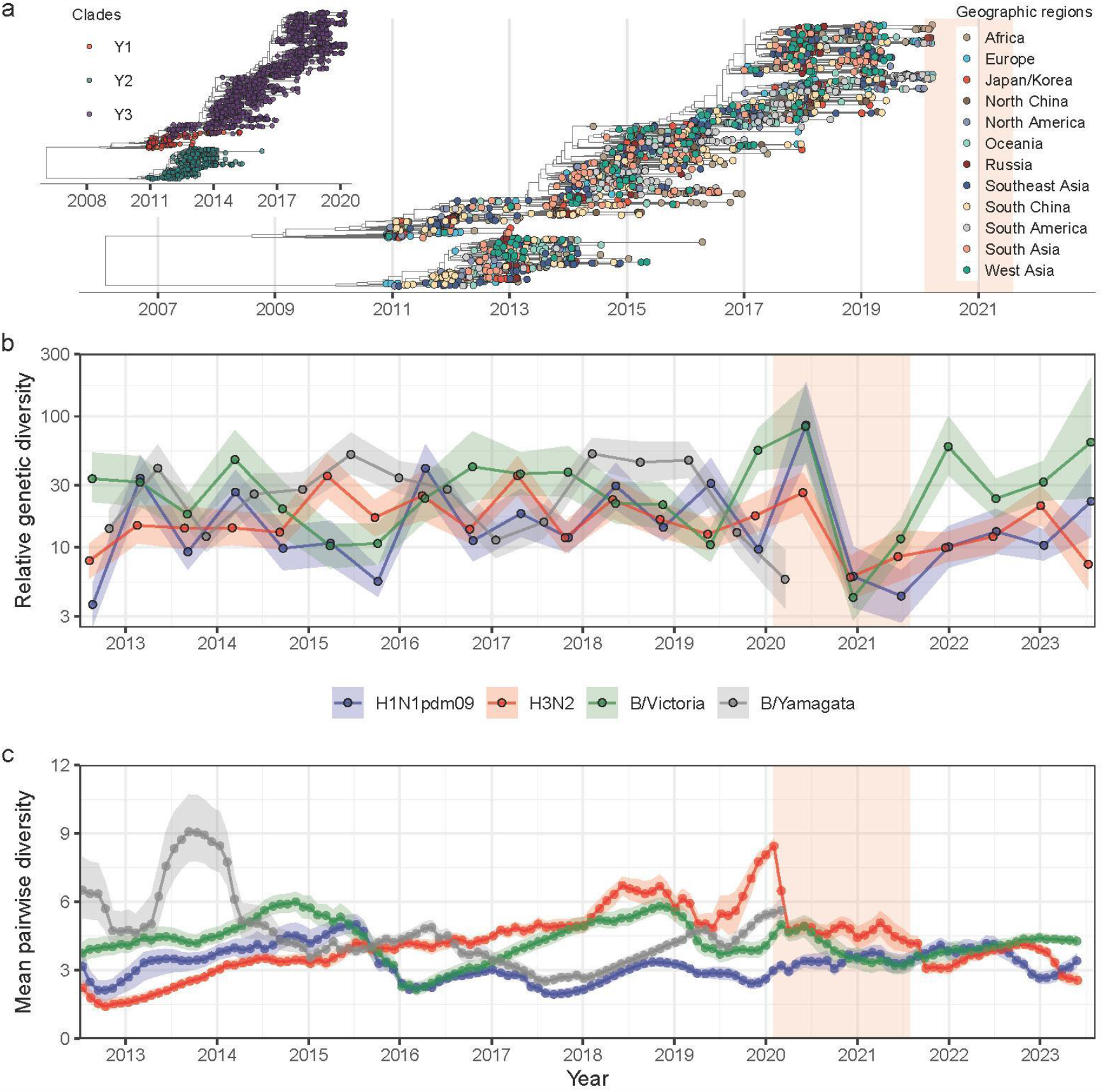
Genetic diversity of seasonal influenza viruses. **a**, Maximum clade credibility (MCC) tree for B/Yamagata lineages. Tip colours represent the geographic regions of the sample. The inset shows the MCC tree with tips annotation representing the main B/Yamagata clades. **b**, Relative genetic diversity of influenza viruses inferred by Bayesian skygrid population reconstruction. **c**, Mean pairwise diversity of influenza virus, measured as average branch length distance in the unit of years between pairs of tips in phylogeny at monthly intervals.

To capture comparative evolutionary dynamics of human influenza virus lineages, we assembled a global genetic dataset of influenza viruses dating back to 2011 (see **Methods**). The relative genetic diversity of influenza A viruses and the B/Victoria lineage declined during the NPIs-intensive phase of the pandemic, and started to accumulate genetic diversity again from the summer of 2021 (**Fig. 5b**). The reduction in genetic diversity was weaker for H3N2 viruses as multiple clades co-circulated (**Extended Data Fig. 10-11**) and accordingly mean pairwise diversity increased since 2013, compared to previous years^29^ (**Fig. 5c**). The relative genetic diversity of the B/Yamagata lineage started to decline in 2019 after peaking in 2018, during a time when B/Victoria genetic diversity was increasing. During 2018-2019, there was only a single clade (Y3 clade) of B/Yamagata lineage remaining, which had been singly circulating since 2015 (**Fig. 5a**). The dominance of the Y3 clade after 2015 was linked to a reduction in mean pairwise diversity of B/Yamagata (**Fig. 5c;** also evidenced in dates of common ancestry, see **Extended Data Fig. 12d**). This suggests that genetically similar B/Yamagata lineages circulated each season after 2015. A lower accumulation rate of nonsynonymous substitutions (d_N_/d_S_: 0.136) and fewer amino acid residues under positive selection (n = 1) in the HA segment of B/Yamagata lineage points to a lower positive selective pressure on the virus (**Supplementary Table 5**), which might correspond to less or slower antigenic drift^30^. In contrast, B/Victoria lineages exhibited a slightly greater d_N_/d_S_ (0.166) and a higher number of amino acid residues under positive selection (n = 4) (**Supplementary Table 5**) in the HA segment that are potentially immune-evasive^24,31^.

The possible “extinction” of B/Yamagata lineages might be explained by a combination of susceptible host depletion due to a large outbreak in 2017-18, NPIs, and slow antigenic evolution^30,32^. More mathematical modelling and empirical investigations are necessary to assess their relative contributions to the possible “extinction” of B/Yamagata.

## Discussion

We find that the NPIs implemented during the COVID-19 pandemic re-shaped the global dissemination of seasonal influenza. During the period of intensive NPIs, influenza transmission declined globally and only in a few geographical regions did we observe substantial lineage movements. Even though the intensity of global human mobility was greatly reduced, international travel remained the principal driver of the international dissemination of seasonal influenza. Lineage transmission persisted more in regions such as West Asia, Southeast Asia, and Africa. Missing data from Africa meant that its role was previously less explored using phylodynamic approaches^15^. Further data is needed to confirm findings about the role of Africa in the seasonal dissemination of influenza^33^. As air travel resumed, so did seasonal influenza transmission, however global virus lineage movements were less intense than before the pandemic. In late 2023, it appears that influenza outbreaks across some parts of the world are more synchronised than in past years, possibly due to higher availability of susceptible populations^34^.

Other studies have documented the transient alteration of global population structure of H3N2 after the air traffic disruptions in Asia Pacific during the SARS outbreak in 2003^35^. During that time, Southeast Asia temporarily acted as the trunk location instead of China^1,15,36^. Our empirical findings from the COVID-19 pandemic consolidate the relationship between the global spread of influenza and changes in global human movement. While it is likely that other factors (climate, demographics, and others) play an important role in global influenza lineage dynamics, they were not the focus of our study. Nonetheless, assessing the relative contribution of each of these factors to seasonal influenza epidemics merits further investigations that might lead to improved intervention strategies and vaccine policies in the future^37^.

In addition to low levels of influenza incidence, substantial virus genetic bottlenecks have also been observed during the pandemic period^8,38^. The B/Yamagata lineage seems to have disappeared after the start of the COVID-19 pandemic and the potential underlying mechanisms for this occurrence have been widely discussed^11,38^. It remains unclear, however, whether B/Yamagata persists below detection levels or is entirely eliminated^39^. Previous studies have highlighted the interplay among population size, virus mutation, and cross-immunity in predicting the circulation of seasonal influenza, and a shift in any or multiple factors might contribute to lineage extinction^32^. The evolutionary and circulation dynamics of seasonal influenza viruses are maintained by a balance between the steady decrease in population susceptibility due to immunity build-up against circulating strains and the emergence of antigenically novel strains. B/Yamagata lineages were characterised by a lower R_e_ and lower levels of antigenic adaptation compared to B/Victoria, potentially limiting its pool of potentially newly susceptible individuals^30,31^. Previous work has shown that extensive genome segment reassortment between B/Victoria and B/Yamagata means that only three segments (HA, PB1, PB2) have an evolutionary history that is specific to either one lineage or the other^28^. If the B/Yamagata lineage has indeed disappeared, monitoring its impact on changing infection rates of B/Victoria should be a priority, as some level of cross-protection immunity is conferred between infections of influenza B viruses^40^.

We interpret our phylodynamic results in the context of several limitations. First, as discussed elsewhere, the inferred number of virus lineage movements do not capture the number of infected individuals due to incomplete sampling and uneven sequencing coverage. We therefore adopted multiple sub-sampling approaches, all of which point towards the same conclusions (**Extended Data Fig. 4, Supplementary Fig. 8**). Second, the three periods selected to perform our analysis are based on air travel data and the COVID-19 stringency index, both of which might not account for all pandemic-related interventions. We therefore performed sensitivity analysis changing the cutoffs defining the third period and our conclusions remain robust to them (**Supplementary Fig. 9**). Third, even though we do not find support for variables other than air traffic in explaining dispersal of seasonal influenza, that might be due to the granularity and scale of our analysis. For example, humidity data were aggregated to macroscopic regions from grid level resolution with high heterogeneity between grids. Evidence from other studies is clear that climatic and urbanisation shape the influenza seasonal influenza transmission^22^. Higher resolution, open, and globally-representative virus genomes, combined with methodologies capable of accommodating tens of thousands of sequences, might address these issues in the future.

Our study provides insights into the impact of the COVID-19 pandemic on vaccination strategies for seasonal influenza. The persistence of seasonal influenza in several regions suggests a delay in the competition among existing influenza strains and the probability of geographically structured viral strains emerging through regional diversification. This could limit universal recommendations for vaccine strain composition for the next season. Generally, seasons with extremely high incidence levels in the post-pandemic period might be associated with vaccine mismatch or the emergence of novel clusters of influenza virus^41^, which requires enhanced surveillance to track the novel virus strains. Except for vaccine-induced immunity, low accumulation of population immunity due to lack of natural infection might likely aggravate future influenza epidemics, as illustrated by the resurged wave in China in the winter of 2023^42^.

The evolutionary and ecological dynamics of influenza globally depend on multiple complex factors, including the transmission potential of strains within locations and the connectedness between locations. The COVID-19 pandemic provided a unique opportunity to evaluate the impact of NPIs on the global circulation of influenza, which has implications on the control and mitigation of future influenza epidemics and pandemics. Our study provides only a short-term, empirical evaluation of the impact of COVID-19-related NPIs, and the longer-term impact of the COVID-19 pandemic on influenza evolution and antigenicity remains unknown^9,43,44^. Our analysis provides a framework for evaluating the impact of pandemics on the global circulation of respiratory infectious diseases beyond SARS-CoV-2 and influenza viruses.

## Methods

### Viral genetic sequence data

We downloaded sequences of the HA segment of seasonal influenza viruses (H1N1pdm09, H3N2, B/Victoria, B/Yamagata) publicly available in GISAID^19^ and NCBI (GenBank)^45^ obtained from human samples on 7 September 2023 (**Supplementary Fig 1**).

Since the genetic data of influenza B viruses deposited in GenBank is not classified into specific lineages, we initially aligned these sequences (reference genome: NC_002207) in Mafft^46^ and constructed a Maximum Likelihood (ML) phylogeny using FastTree v2.1.11 under a general time-reversible (GTR) substitution model to classify them into Victoria and Yamagata linages^47^. For each seasonal influenza virus, we performed a sequence alignment and quality assessment using the Nextclade pipeline where we only retained genetic data with good quality defined by the quality control criteria in Nextclade^48^. Since the analyses would be based on the coding region of sequences, we kept the sequences with over 984 bp in coding region^15^. We further removed genetic data lacking complete information about collection dates or locations, and here we only focused on genetic data from geographic locations within WHO member states.

### De-duplication of genetic data

Multiple specimens from a single individual patient can sometimes be uploaded to a single database; furthermore, the same sequence is sometimes uploaded to both GenBank and GISAID. We therefore implemented a de-duplication procedure applied to sequences within and between repositories. Within each repository, we identified and removed duplicates based on the same isolate names, retaining the one with the earlier collection date (with first priority) and higher genetic coverage. After this initial step, genetic sequences were queried amongst both repositories and considered as duplicates if 1) they have the same isolate names or; 2) have the same sampling location (level-0 boundary, country level), collection date, and raw genetic sequence. Technically, the second rule cannot completely assert that they are duplicates, while we introduced it since the entry criteria of isolate names might vary across repositories and we aimed to minimise the likelihood of including unidentified duplicate samples. The above curation processes assembled a set of high-quality genetic dataset.

Next, based on the spatial proximity and potential roles in disseminating influenza^15^, we discretised the sample collection locations by clustering them into 12 distinct geographic regions: North America (comprising Canada and the USA only), South America, Europe, Russia, Africa, West Asia, South Asia, North China, South China, Japan/Korea, Southeast Asia, and Oceania (**Supplementary Fig 2**). Sequences from Central America, Central Asia, and parts of East Asia (countries other than Japan, Korea and China) were excluded from subsequent phylogenetic and phylogeographic analyses due to limited number of available genetic data and their potentially less significant role in disseminating seasonal influenza^15^. Overall flowchart of collating genetic data was presented in **Supplementary Fig 1**.

### Epidemiological surveillance of seasonal influenza

Weekly notified cases of seasonal influenza and specimens to be processed for influenza testing at the country/territory level were downloaded from the FluNet tool on 7 September 2023, which are provided remotely by the Global Influenza Surveillance and Response System (GISRS)^16^. We excluded records considered to be uninformative or unreasonable due to numerical inconsistencies (e.g., the combined total of positive influenza A, positive influenza B and negative specimens greatly exceeded (here set to 5) the total number of processed specimens, which has not been set equal given the potential co-infection of influenza A and B^49^), with a total of over 30,000 rows removed. We imputed the unsubtyped, unsubtypable or lineage-undetermined samples into specific subtypes or lineages based on the available weekly- and country-specific proportion of subtypes or lineages, in which a Bayesian framework with uninformative Beta (1, 1) prior was used to calculate the posterior proportions and 95% uncertainty levels given that small tested size might cause extreme scaling.

Subsequently, we summarised the number of specimens to be processed for influenza testing over time to reflect the intensity of virological surveillance and computed the rolling positivity rate (5-week window) of each influenza subtypes/lineages among all tested specimens on a weekly basis, with interval estimates determined by the 95% uncertainty levels for inferring the specific subtype or lineage. Intensity of genomic surveillance of influenza was indicated by the rolling percentages of reported lab-confirmed influenza cases being sequenced. To minimise the impact of heterogeneous surveillance intensity across periods, we also estimated the transmission activity level via calculating the ratio of positive influenza cases to the total number of specimens processed in each period (defined later).

### Genetic sampling and selection

Given that the phylodynamic inference is sensitive to sampling bias^27,50,51^, we carefully selected and sub-sampled genetic sequences in order to make their distributions as representative/even as possible while enhancing computational efficiency. In phylogeographic analyses, we only focused on the most recently circulating sub-clades of each seasonal influenza virus. Specifically, we chose the 6B.1A.5 sub-clade and its descendants for H1N1pdm09, 3C.2a1b sub-clade and its descendants for H3N2, V1A.3 sub-clade and its descendants for B/Victoria, and Y3 clade with M251V amino acid mutation for B/Yamagata.

To mitigate the impact of surveillance biases, we proposed an even sampling strategy, similar to^15^. Briefly, we evenly subsampled the sequences across 12 geographic regions and years via precisely defining the number of sequences to be randomly subsampled per region per year (ensuring the total sub-sampled numbers of genetic sequences are about 2,000), which would ensure an equitable distribution across years and geographic regions for each influenza subtype/lineage^15^. Additional genetic data were randomly selected where available to ensure that there was at least one sequence sample per week throughout the evaluation period.

Besides, we also adopted two additional sub-sampling strategies, namely, strategy accounting for temporal variation and strategy accounting for spatiotemporal variation. In both schemes, we first determined the weekly number of sequences to be subsampled at the global level in direct proportion to the product of the global positivity rate of each seasonal influenza virus and the proportion of recently circulating clades that we selected for analyses. With these weekly numbers in hand, sequences were evenly sampled across 12 geographic regions for each week for the former scheme; sequences were sampled with a weight based on the counts of subtype/lineage-specific influenza cases per geographic region per week in the latter scheme, with the guarantees of approximately 2,000 genetic sequences selected as well. For weeks with particularly low influenza positivity rates, at least 3 sequences were sampled where possible to maintain consistency in the temporal scale.

To capture more evolutionary dynamics in the past, we additionally adopted the even sub-sampling strategy to assemble a global genetic dataset with about 2,000 sequences dating back to 2011.

### Phylogenetic inference

First, we constructed maximum likelihood (ML) phylogenies for the above sub-sampled genetic data using IQ-TREE v2.0.7^52^ under the nucleotide substitution model automatically selected by ModelFinder^53^. The resulting phylogenetic trees were inspected with best fits in TempEst v1.5.3 to identify and remove temporal outliers^54^, defined as when residuals deviated by more than three standard deviations from the mean. We then inferred the time-calibrated tree using TreeTime v0.11.1^55^, which would serve as the starting tree for the Bayesian phylogenetic inference.

Phylogenetic trees were inferred for H1N1pdm09, H3N2, B/Victoria and B/Yamagata in Bayesian framework using BEAST v1.10.4^56^, in which we incorporated a starting ML tree, a SRD06 nucleotide substitution model^57^, a Bayesian SkyGrid coalescent prior (with grid points equidistantly spaced in six month intervals)^58^, and a strict molecular clock model with a lognormal distribution prior. Markov Chain Monte Carlo (MCMC) was run in parallel for 2 independent chains, with a total of about 250 million steps sampled every 50,000 steps for each chain. Steps were combined across two chains with the first 10% discarded as burn-in. MCMC convergence was checked in Tracer v.1.7.1^59^ and effective sample sizes (ESS) for all continuous parameters are over 100. We resampled states every 400,000 steps, which yields a total of approximately 1000 empirical trees for each influenza subtype/lineage under each sub-sampling strategy.

### Time-inhomogeneous phylogeographic reconstruction

We used the empirical tree distributions to further perform the phylogeographic reconstruction under the generalised linear model (GLM) framework^1^. The default model assumption with constant-through-time rate of viral geographic dispersal along the phylogeny might not fit the scenario where the adoption of NPIs against the COVID-19 pandemic may alter the transition rate of other respiratory infectious diseases. Therefore, we adopted a time-inhomogeneous GLM model to allow for incorporating interval-specific predictors and accounting for the potential variation in rate of influenza dispersal before, during, and after the NPIs-intensive phase of COVID-19 pandemic^25^. The cutoff points that defined three phases were 1) 31 January 2020, in which the global COVID-19 stringency index increased rapidly and global air traffic volume started to decline, and 2) 31 July 2021, when the global COVID-19 stringency index gradually declined, global air traffic volume started to resume, and the positivity rate of seasonal influenza started to increase (**Fig 1, Extended Data Fig. 1**).

Specifically, we first adopted a three-period GLM-diffusion phylogeographic model with interval-specific indicator variables to represent the inclusion or exclusion of predictors^25^. Predictor inclusion was further pooled across intervals using hierarchical graph modelling to allow for including covariates at the hierarchical level^60^. Hierarchical and interval-specific indicators were estimated under spike-and-slab procedure^60^. Here, multiple categories of potential predictors of spatial spread of influenza (e.g., demographic, meteorological, air passenger traffic, geographic, and sampling factors) were included in the GLM model (**Supplementary Fig 3-5, Supplementary Table 1-3**). Among that, air traffic data were provided by the Official Airline Guide (OAG), with details in the **Appendix**. Notably, we only adopted a time-homogeneous (1-period) model for B/Yamagata given the limited sequences sampled after the onset of COVID-19 pandemic.

Given that air traffic between regions is able to significantly predict the spatial spread of influenza indicated by the above analyses (**Fig. 2g**), we subsequently specified a three-period GLM model with air traffic data as sole predictor for the relative rates and extended it to also include overall air traffic as a predictor for the overall transition rate scalar. Specifically, we incorporated the log-transformed and standardised average volume of air traffic in each period as a predictor for the overall average migration rate across three periods. In addition, we included the asymmetric air traffic matrix between regions as the predictor for relative transition rates between each pair of regions. Therefore, both the relative and average migration rate are able to vary across the three periods, exactly rationalising the potential impact of COVID-19 pandemic in spatial transmission of influenza. To detect potential deviations in the predictive power of air mobility for specific transition rates, time-homogeneous random effects were added to the parameterization of phylogeographical transition rate^25^. For each influenza subtypes/lineages, a total of less than 6 random effects of transition rate with their 95% highest posterior density (HPD) intervals deviated zero, indicating a good predictive ability (**Supplementary Fig 12**). The complete Markov jump history through time was also inferred under this model. A total of 10 million steps were run for at least one chain and sampled every 5,000 steps in GLM phylogeographic models.

### Summary of posterior trees

First, we summarised the trunk location through time based on the phylogeographic estimates using PACT v.0.9.5 (https://github.com/trvrb/PACT)^15,36^, where the trunk is defined as all branches ancestral to viruses sampled within 1 year of the most recent sample. Second, we adopted the TreeMarkovJumpHistoryAnalyzer tool to obtain the posterior summaries of Markov jumps and their timings. Upon this, we inferred the patterns of transition flow and mapped the migration dynamic across the three periods, in which we also calculated the net export during each period. To account for the heterogeneity of lifting time of NPIs across regions, we further adopted the region-specific cutoffs to define the third period based on the region-specific recovery of air traffic (**Fig. 2d**), where 1 December 2021 was used for South Asia; 1 July 2022 for Japan/Korea, Oceania, and Southeast Asia; and 1 December 2022 for China. Third, two metrics of viral persistence were inferred: 1) tip-associated persistence, measured as the time for tips to leave its sampling location walking backwards in time across the trees using PACT v.0.9.5; 2) lineage-associated persistence, measured as the mean time for lineages circulating along phylogenetic branches at a given evaluation time to leave current geographic location through walking backwards across the trees using the PersistenceSummarizer tool. Here, we combined South China and North China together to estimate the persistence in China. Fourth, we estimated the time to the most recent common ancestor of tips circulating per season per hemisphere across trees using the reassembled genetic dataset, as a proxy of lineage turnover. Fifth, mean pairwise diversity (or genealogical diversity) through time was estimated by averaging the pairwise distance in time between each pair of tips with a 1-month window across trees using PACT v.0.9.5.

### Selection pressure

Selection pressures for the HA segment of each influenza subtypes or lineages were inferred as the ratio of the number of nonsynonymous substitutions per nonsynonymous site (*d*_*N*_) to the number of synonymous substitutions per synonymous site (*d*_*S*_) using the methods of Single Likelihood Ancestor Counting (SLAC)^61^ and Fast Unconstrained Bayesian AppRoximation (FUBAR)^62^ in the HyPhy package^63^. Amino acid sites were considered to be under positive or negative selection if they were selected by the two methods, where *p*-value threshold of 0.1 in SLAC and posterior probability of 0.8 in FUBAR were used to identify those sites under selection.

### Bayesian hierarchical regression model

In what follows, we explore the potential drivers of dynamic patterns of lineage-associated persistence over time. Previous work indicates that local persistence of influenza virus is associated with antigenic drift and seasonality^15^, from which we further hypothesise that NPIs adopted during the pandemic, in particular the reduction of inter-region air traffic arrivals, might contribute to persistence. Here, we constructed a Bayesian hierarchical regression model to model the association of antigenic drift (*w*) and air traffic arrival reductions (*x*) with lineage-associated persistence (*y*) (here measured in the units of years and changed every month) while accounting for regions and months (conceptual details presented in **Supplementary Fig 13**).

### Standardisation of variables

We first use genetic data to measure cross-immunity to indicate the extent of antigenic drift^26,64^. Briefly, for each virus strain *i* in a three-month time window (e.g., from Jan 2020 to Mar 2020), we limited pairwise comparisons to sub-sampled global strains *j* circulating within the last six months (e.g., from Jul 2019 to Dec 2019) from the current three-month time window (**Supplementary Fig 14**). We counted the number of amino acid differences between the HA segments at 49 epitope sites for each pair of strains in adjacent time windows^26^. Then we estimated the ability of strain *i* to escape immunity by summing the mean exponentially-scaled epitope distances between the given strain (*i*) and previously circulating strains (*j*) using^26^:

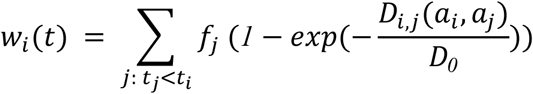

Where *i* is the given strain in the current three-month time window (*t*_*i*_, e.g., from Jan 2020 to Mar 2020), *j* are the strains circulating in the previous six-month time window (*t*_*j*_, e.g., from Jul 2019 to Dec 2020), *f*_*j*_ is the strain frequency in the previous time window, *D*_*i,j*_ is the epitope distances between strain *i* and strain *j*, and *D*_*0*_ is the constant (set as 14)^26^. Since each virus strain *i* was associated with a region *r* and a three-month period *m*, we summarised the average value (*w*_*r,m*_) by region and three-month time window. We then standardised these region- and month-specific antigenic drift *w*_*r,m*_ as follows:

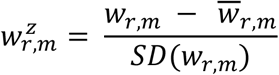

Where *w*_*r,m*_ is the antigenic drift of strains circulating in region *r* in month *m* (constant through three months, e.g., *w*_*r,Jan*_ = *w*_*r,Feb*_= *w*_*r,Mar*_), and 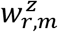 is the z-score of standardised *w*_*r,m*_, namely *w*_*r,m*_ minus its mean 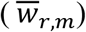 divided by its standard deviation (SD).

For each month *m* in region *r*, we calculated *x*_*r,m*_, defined as the relative reduction of inter-region air traffic arrivals (*A*_*r,m*_) compared to the monthly maximum:

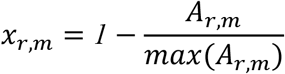

And we then computed the standardised z-score 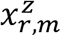 :

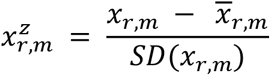

We also standardised the persistence (*y*_*r,m*_) as the outcome/dependent variable:

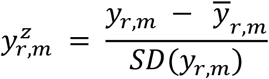

Where *y*_*r,m*_ is the lineage-associated persistence of viruses circulating in region *r* in month *m* (evaluated as the midpoint of each month), and 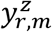 is the z-score of standardised *y*_*r,m*_.

### Regression model

Afterwards, we model the association of standardised antigenic drift 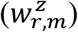 and standardised air traffic reduction 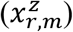 with standardised persistence 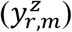 in a Bayesian hierarchical model. According to the directed acyclic graph (DAG) framework we assumed (**Supplementary Fig 13**), we first predicted antigenic drift as normally-distributed observations with mean γ that is a linear function of one predictor variable 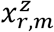 :

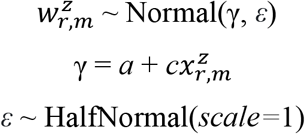

Where *a* is the intercept, *c* is the coefficient for covariate 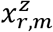, while *ε* represents a half-normal-distributed observation error.

We then predicted persistence as Gaussian-distributed observations with mean *μ* that is a linear function of two predictor variables, 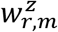 and 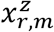 :

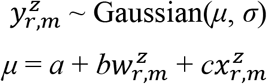

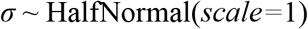

Where *a* is the intercept, *b* is the coefficient for covariate 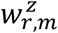, and *c* is the coefficient for covariate 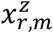, while *σ* represents a half-normal-distributed observation error. This linear sub-model is linked to the previous one via *a* and *c* parameters.

Since we are constructing a Bayesian model, we should assign prior distributions to the unknown *a, b*, and *c*. To facilitate MCMC sampling, we structured these parameters (*a, b*, and *c*, same as described above) in a non-centered parametrisation. We use a half-normal distribution (normal distribution folded at zero) as the prior for scale parameters (*a*_*s*_, *b*_*s*_, *c*_*s*_), and standard Normal priors (i.e. mean = 0, sd = 1) for location parameters (*a*_*l*_, *b*_*l*_, *c*_*l*_) and standard Normal distributions for base distributions (*a*_*z*_, *b*_*z*_, *c*_*z*_) with two dimensions: region and month. The non-centered Normal distributions are then derived as the location plus the product of the scale and base distribution (e.g., *a*_l_ + *a*_s_*a*_z_):

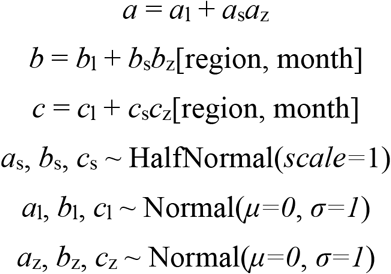

We performed the analyses focusing on the H3N2 virus subtypes circulating in Africa, South Asia, and Southeast Asia. This selection was motivated by the occurrence of local H3N2 epidemic waves in these regions during the NPIs-intensive phase of COVID-19 pandemic (**Fig. 2f**). This allows for estimating the lineage-associated persistence through time because more genetic data is available in the second period.

The model was implemented and sampled with PyMC v.5^65^ using a Hamiltonian Monte Carlo (HMC) No-U-Turn Sampler (NUTS) with four parallel chains of 2,000 tuning steps, 2,000 samples, and a target acceptance probability of 0.99. The chains mixed well with all bulk effective sample size (ESS) > 1000, all Gelman-Rubin convergence diagnostic 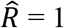, and all Bayesian Fractions of Missing Information (BFMIs) >= 0.75.

## Data and code availability

Genetic sequences used were available in NCBI (https://www.ncbi.nlm.nih.gov/labs/virus/vssi/#/) and GISAID (https://www.gisaid.org/). Influenza virological surveillance data were available from the FluNet (https://www.who.int/tools/flunet). The origin-destination passenger air traffic data up to July 2023 were provided by Official Airline Guide (OAG) Ltd. (https://www.oag.com/) through a data sharing agreement. The codes and accession IDs of sequences used to run the phylogenetic analysis are available here: https://github.com/zychenfd/global_influenza_project.

## Data Availability

The codes and accession IDs of sequences used to run the phylogenetic analysis are available here: https://github.com/zychenfd/global_influenza_project.

## Acknowledgements

We gratefully acknowledge all data contributors, i.e., the Authors and their Originating laboratories responsible for obtaining the specimens, and their Submitting laboratories for generating the genetic sequence and metadata and sharing via the GISAID Initiative and NCBI, on which this research is based. The acknowledgment table of genetic data used is provided on our GitHub repository. We thank Dr. William Wint and Dr. John Brittain for valuable discussions. H.Y. acknowledges financial support from the Key Program of the National Natural Science Foundation of China (No. 82130093) and the General Program of the National Natural Science Foundation of China (No. 82073613). M.U.G.K. acknowledges funding from The Rockefeller Foundation, Google.org, the Oxford Martin School Pandemic Genomics programme (also O.G.P.), European Union Horizon 2020 project MOOD (#874850), The John Fell Fund, a Branco Weiss Fellowship and Wellcome Trust grants 225288/Z/22/Z and 226052/Z/22/Z. P.L. acknowledges support from the European Research Council (grant agreement no. 725422 – ReservoirDOCS). J.C. acknowledges financial support from the Young Scientists Fund of the National Natural Science Foundation of China (No. 82304199).

## Author contributions

M.U.G.K. and H.Y. conceived and planned the research. Z.C., M.U.G.K., P.L., and S.B.M. analysed the data. J.L.H.T., B.G., L.D.P., X.D., J.C., S.B., M.A.S., O.G.P., P.L., M.U.G.K. and H.Y. advised on methodologies. Z.C. and M.U.G.K. wrote the initial manuscript draft. All authors edited, read, and approved the manuscript.

## Extended Data

**Extended Data Fig. 1.**
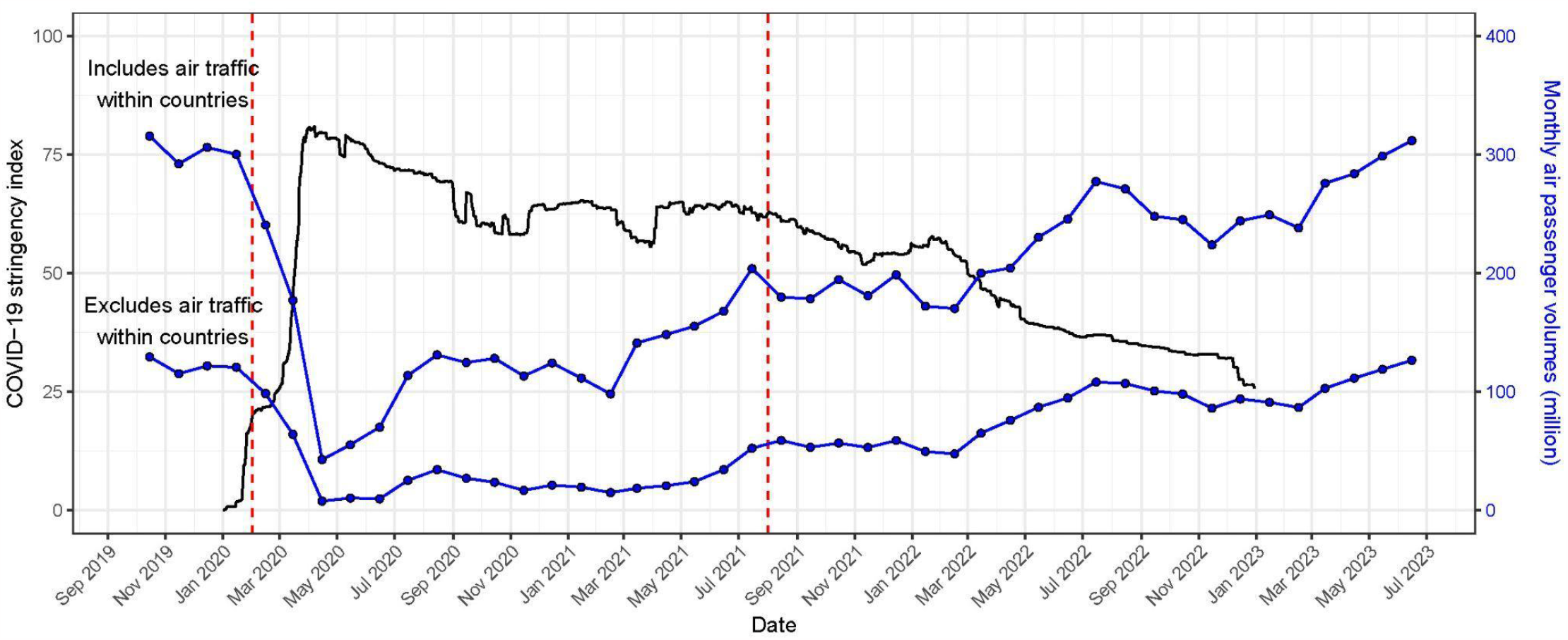
The Global COVID-19 stringency index and monthly air passenger volumes over time. Data for the stringency index were derived from Our World in Data^66,67^, which was weighted by country-level population size to calculate the global index.

**Extended Data Fig. 2.**
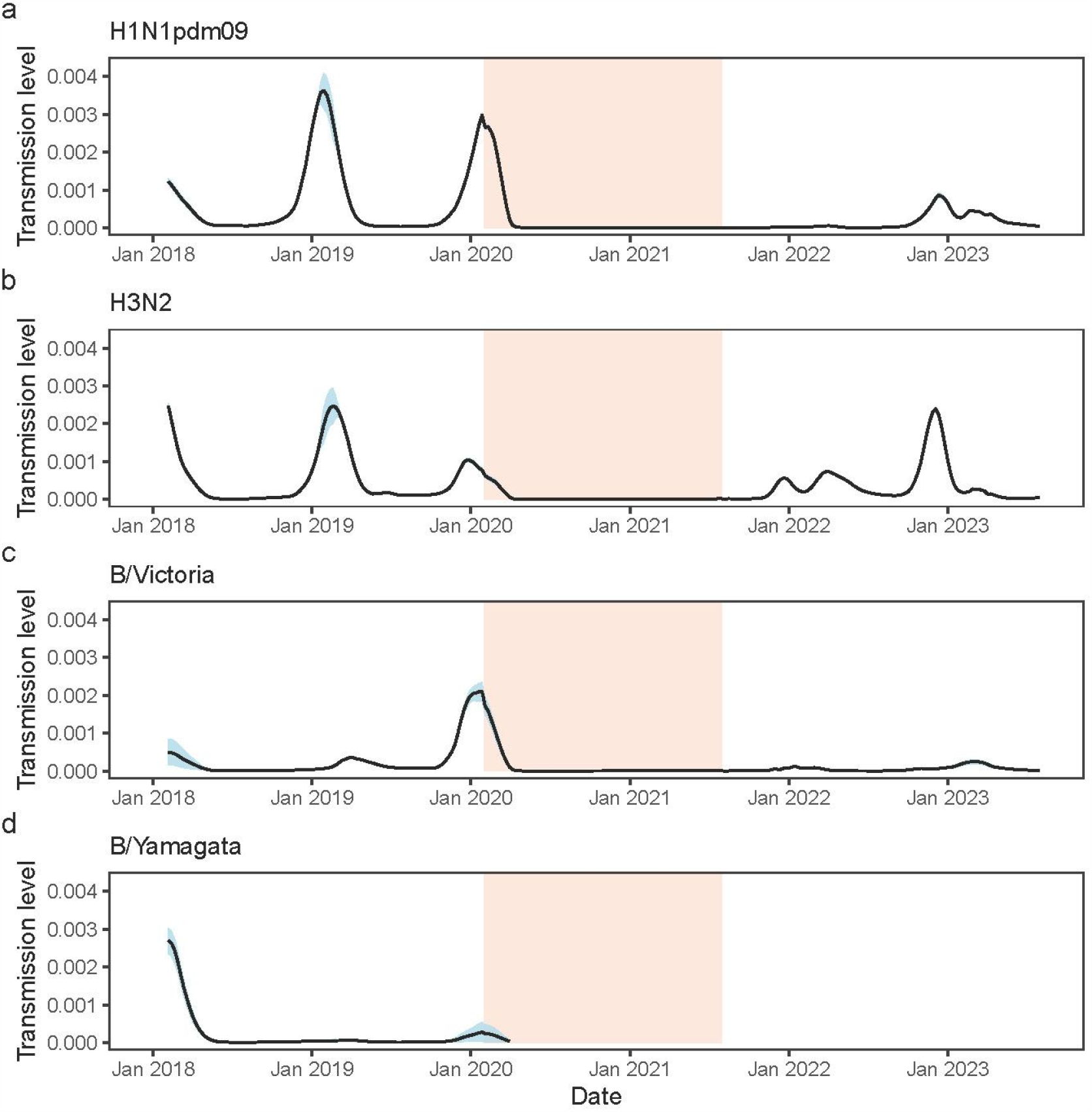
Transmission activity level of seasonal influenza virus accounting for the variations in surveillance intensity across periods. To minimise the impact of variations on surveillance intensity across periods, we estimated the transmission activity level via calculating the ratio of positive influenza cases to the total number of specimens processed in each period. The estimates of absolute numbers are not meaningful, while comparisons over time can be used for the signal of changes in transmission level. The light orange shadow area represents the NPIs-intensive phase of the COVID-19 pandemic (second period), defined as spanning from 1 February 2020 to 31 July 2021.

**Extended Data Fig. 3.**
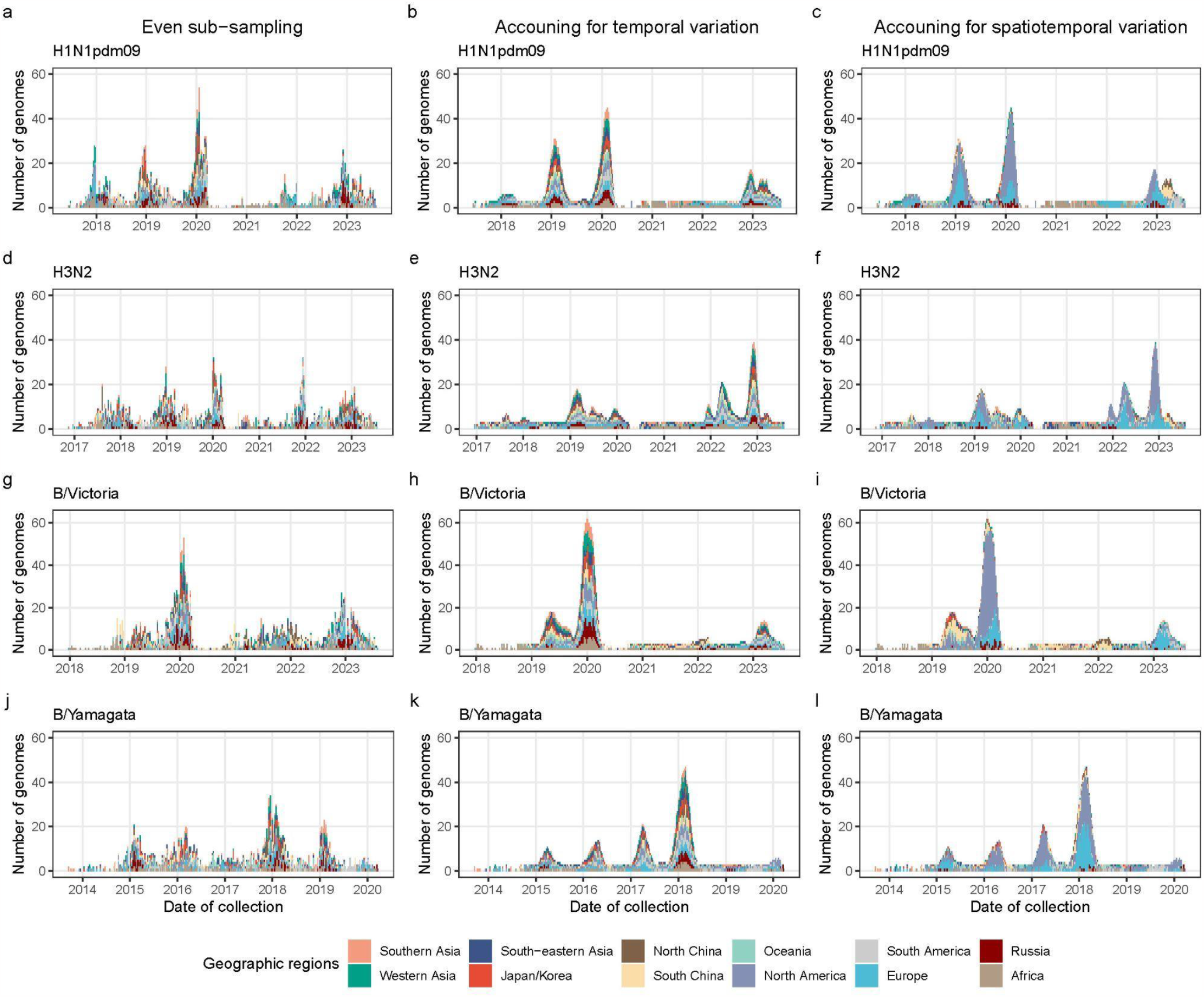
Spatiotemporal distribution of approximately 2,000 genetic data for each influenza subtype/lineage subsampled through three sub-sampling schemes.

**Extended Data Fig. 4.**
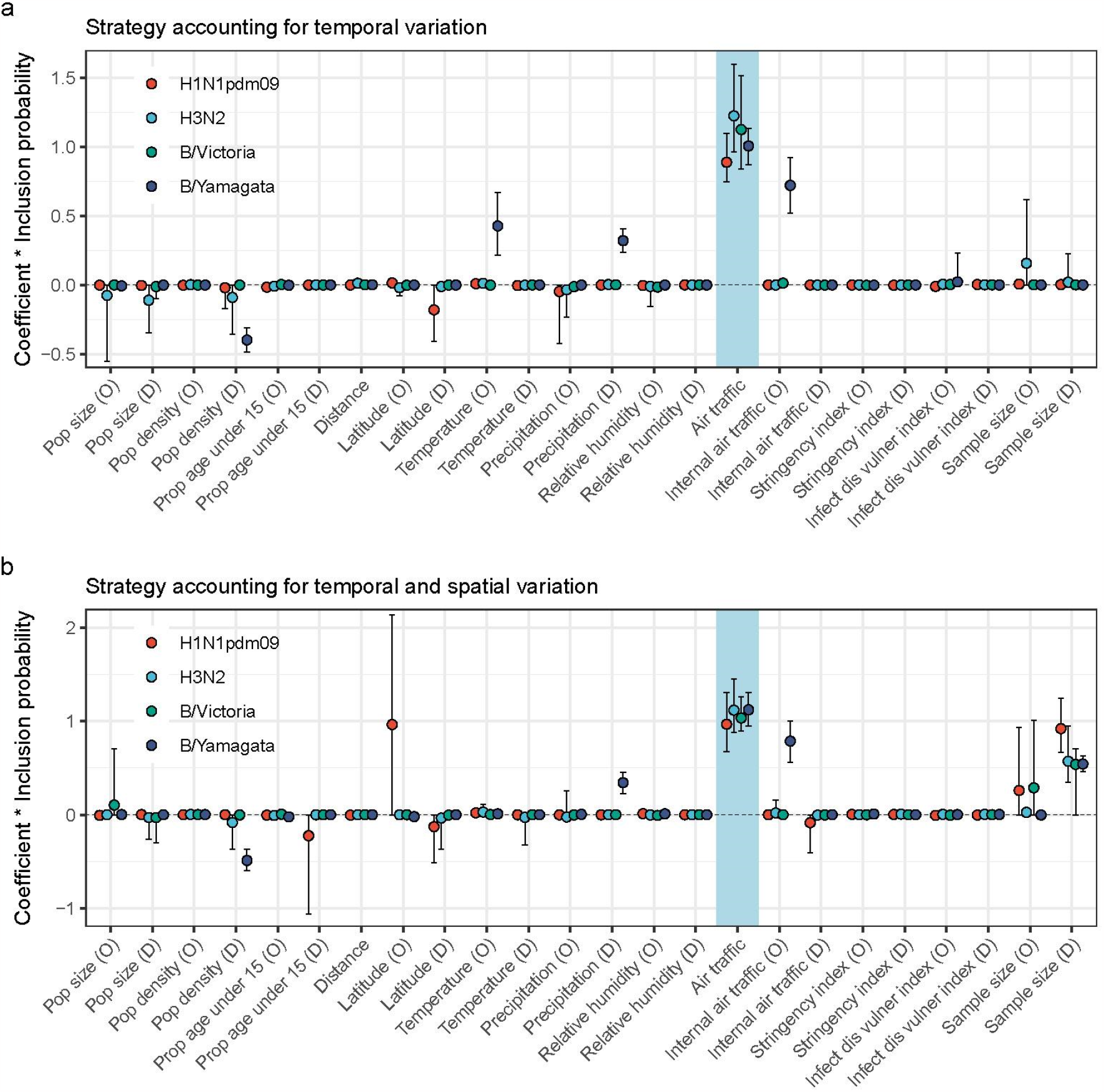
Predictors of global movements of seasonal influenza virus using 3-period GLM model under various sub-sampling strategies. Analyses for B/Yamagata virus were performed under a time-homogeneous (1-period) GLM model since few genetic data available after March 2020. Points and ranges represent the posterior mean and 95% highest posterior density (HPD) intervals, respectively.

**Extended Data Fig. 5.**
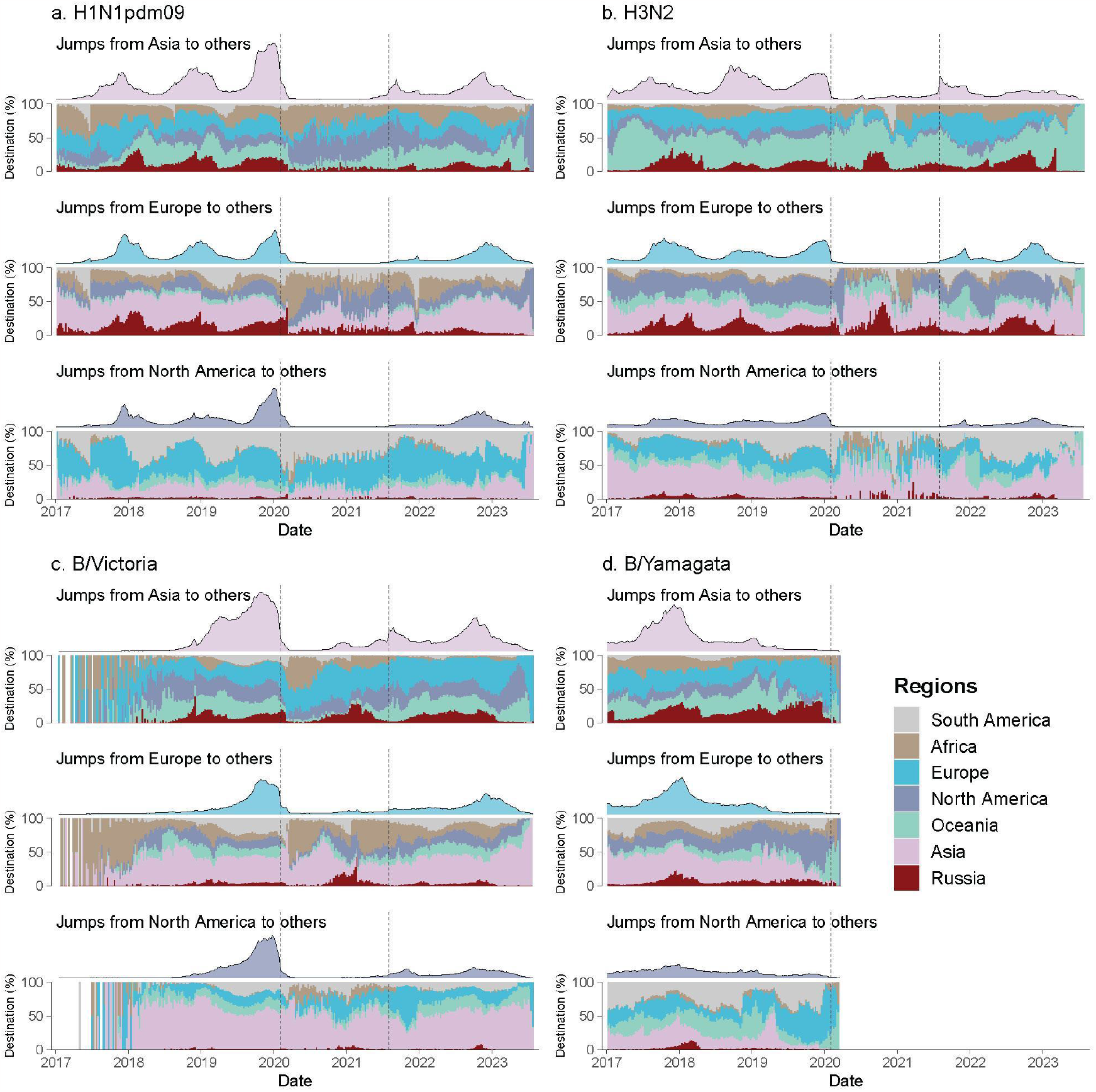
Posterior estimates of weekly counts of location transition from specific regions over time. The curve represents the counts of Markov jumps from each region to other regions over weeks, which was comparable within and across panels. The proportion below was the proportion of destinations of the Markov jumps from specific regions. Notably, proportion was unavailable in some weeks without Markov jump events, and unstable in weeks with few Markov jump events especially during the NPIs-intensive phase of the pandemic. The dashed lines refer to the cutoff points that define the three periods.

**Extended Data Fig. 6.**
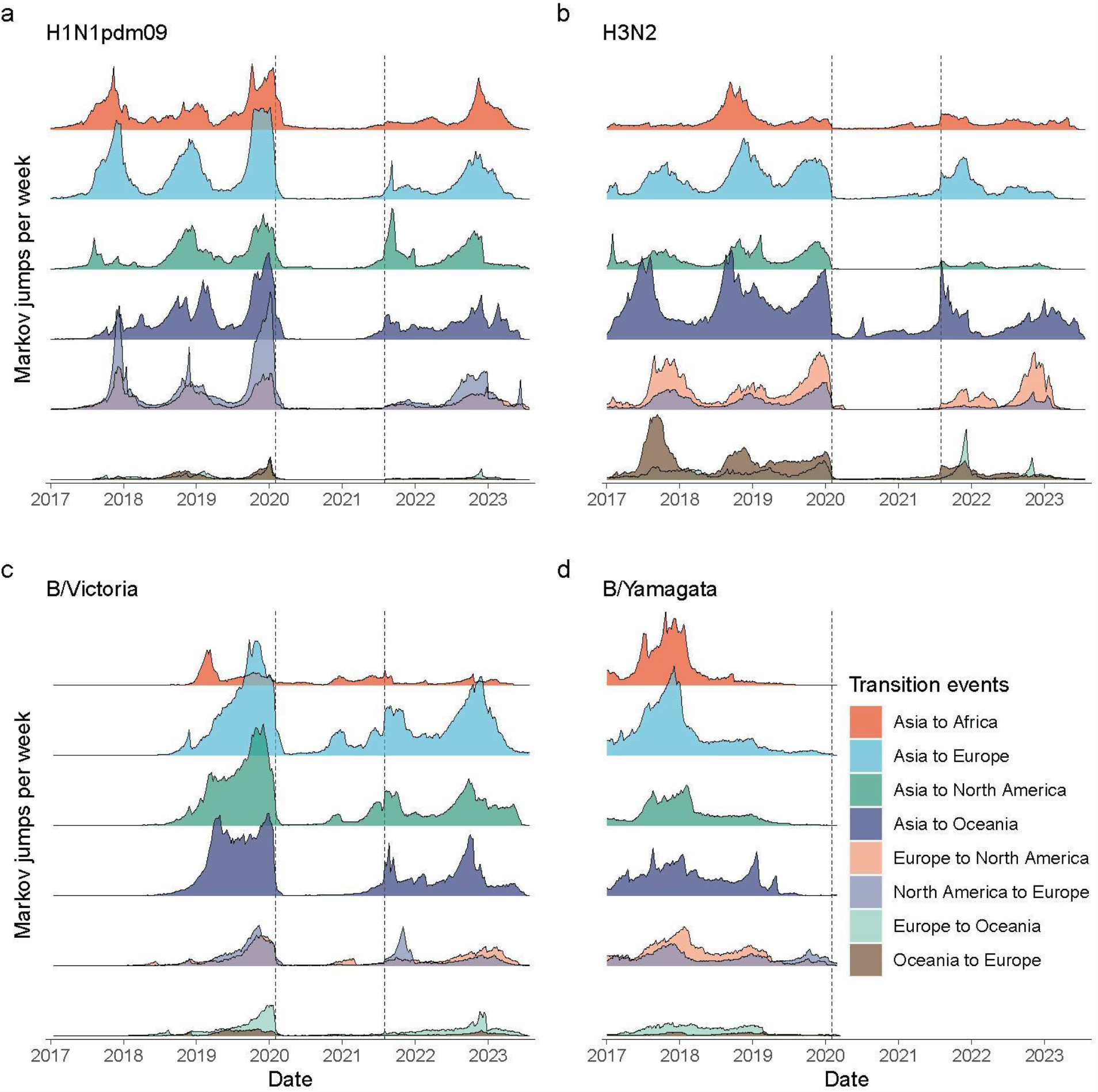
Posterior estimates of weekly counts of location transition between regions over time. The density of Markov jumps was comparable within and across panels. The dashed lines refer to the cutoff points that define the three periods.

**Extended Data Fig. 7.**
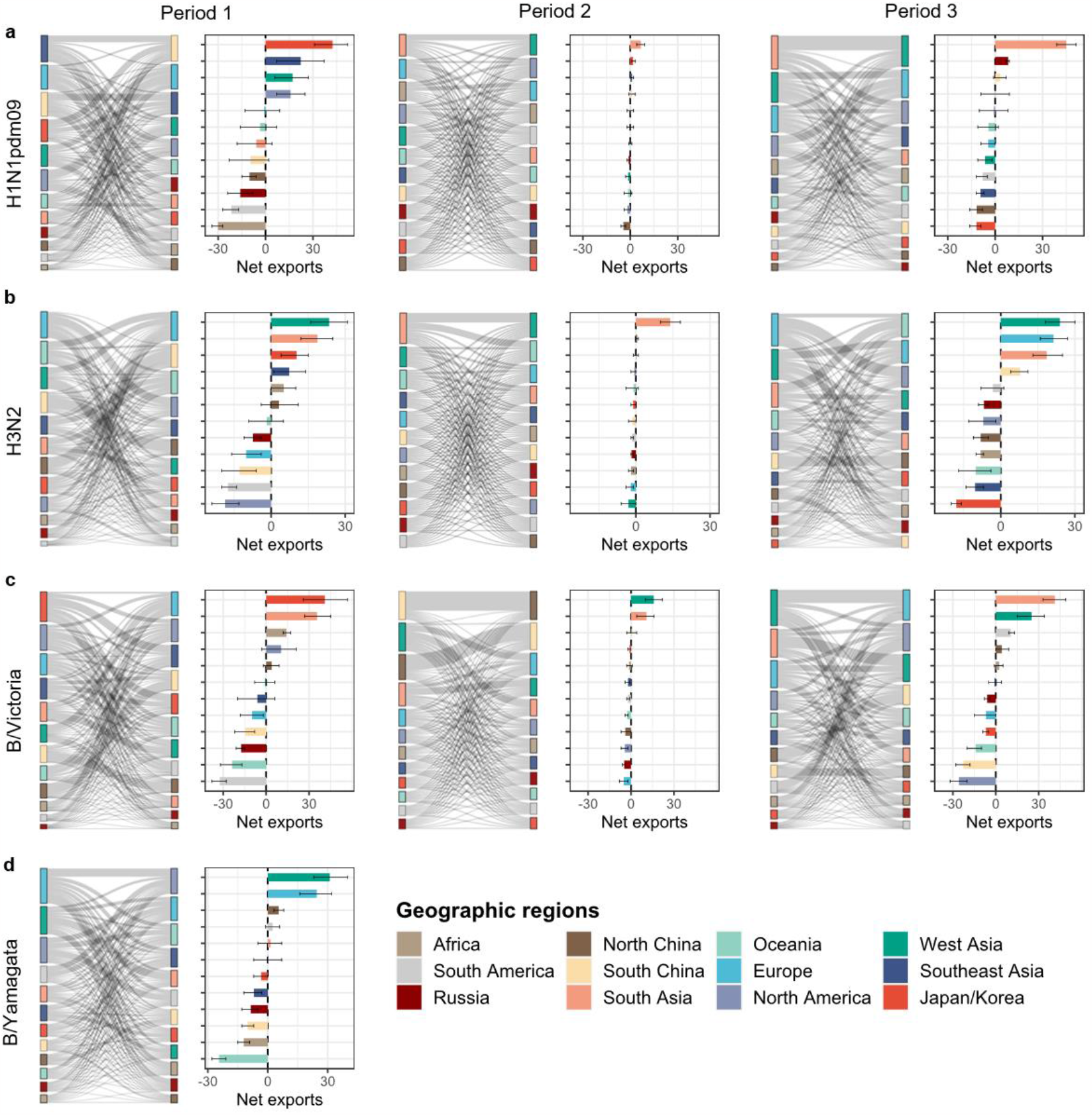
The period-specific net export dynamic of seasonal influenza. Here the first period is defined as periods from February 2018 to January 2020 (24 months); the second period ranges from February 2020 to July 2021 (18 months); and the third period is from August 2021 to July 2023 (24 months). Analyses are based on the posterior summaries of the Markov jumps under a time-inhomogeneous (3-period) GLM model with only air traffic data as the predictor of overall and relative transition rates, except for B/Yamagata under a time-homogeneous (1-period) GLM model. Error bars in the bar charts represent the interquartile range.

**Extended Data Fig. 8.**
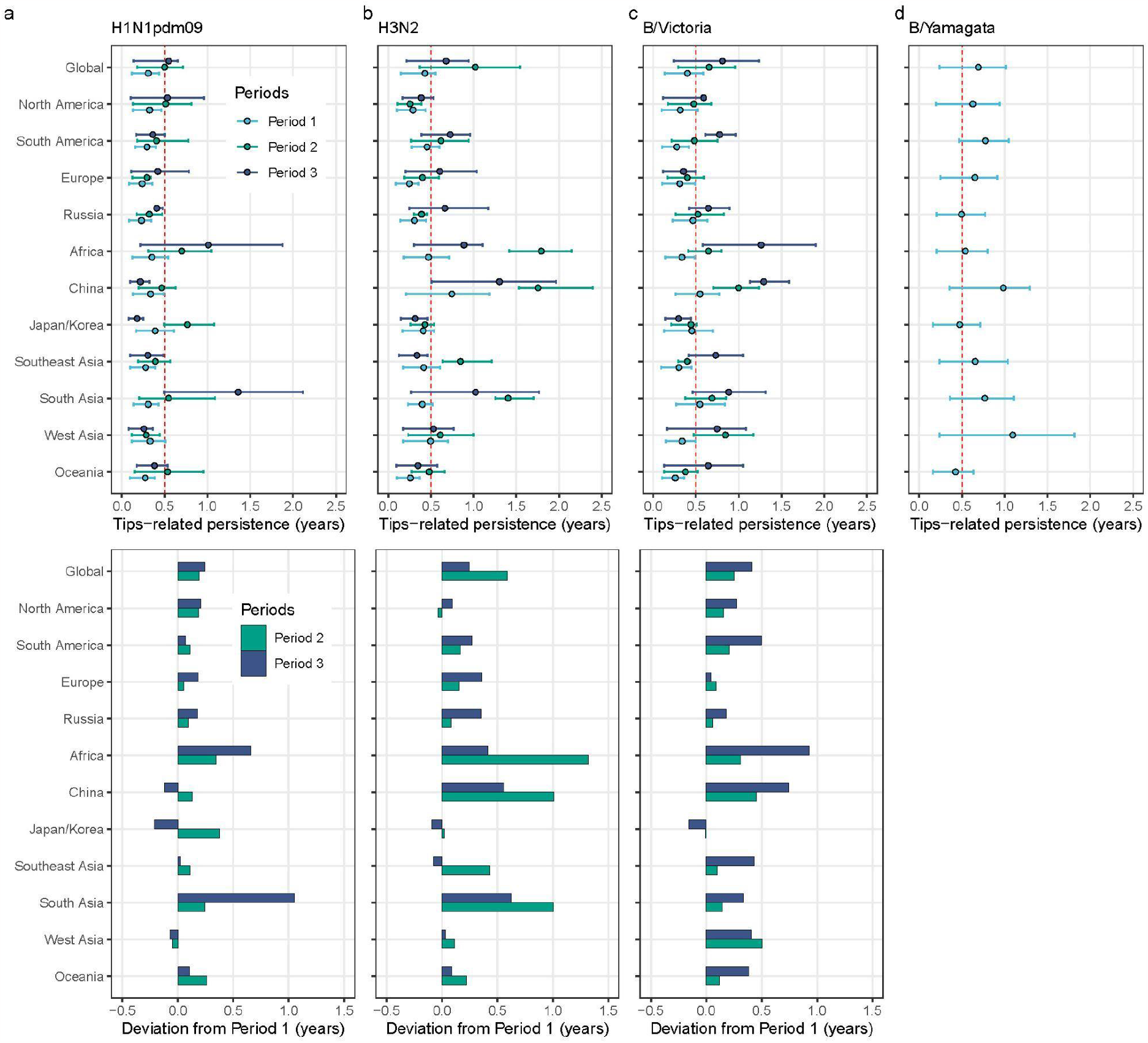
Tip-related persistence across three periods. **a-d**, The persistence of tips for **a**) H1N1pdm09, **b**) H3N2, **c**) B/Victoria across three periods, and **d**) B/Yamagata in the first period. At the bottom panels, we presented the persistence deviations from Period 1. Here we combined North China and South China together to estimate the persistence in China. Tip-associated persistence represents the time for tips to leave its current sampling location through walking backwards up the phylogeny. Points refer to mean persistence and lines represent the interquartile range.

**Extended Data Fig. 9.**
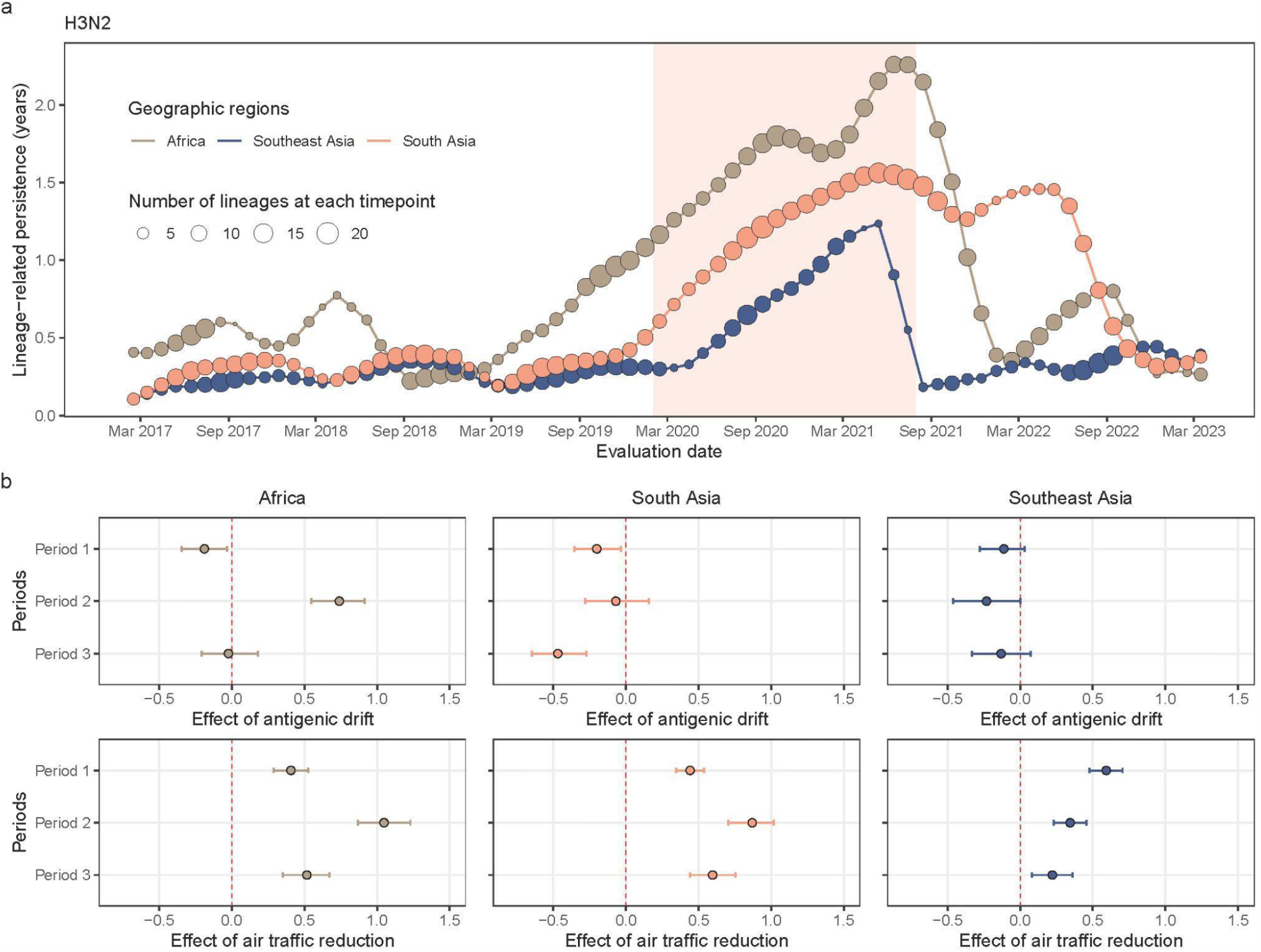
Lineage-related persistence of H3N2 in Africa, Southeast Asia, and South Asia. **a**, The temporal patterns of lineage-associated persistence of H3N2 virus subtypes circulating along the branch at various evaluation times in Africa, Southeast Asia and South Asia. Lineage-associated persistence refers to the mean time for lineages circulating along phylogenetic branches at a given evaluation time to leave current geographic location. **b**, The effect (z-score) of antigenic drift and inter-region air traffic reduction on lineage-associated persistence of H3N2 virus circulating in Africa, Southeast Asia and South Asia in each period, respectively. Points refer to mean estimates and lines represent the 90% highest density intervals.

**Extended Data Fig. 10.**
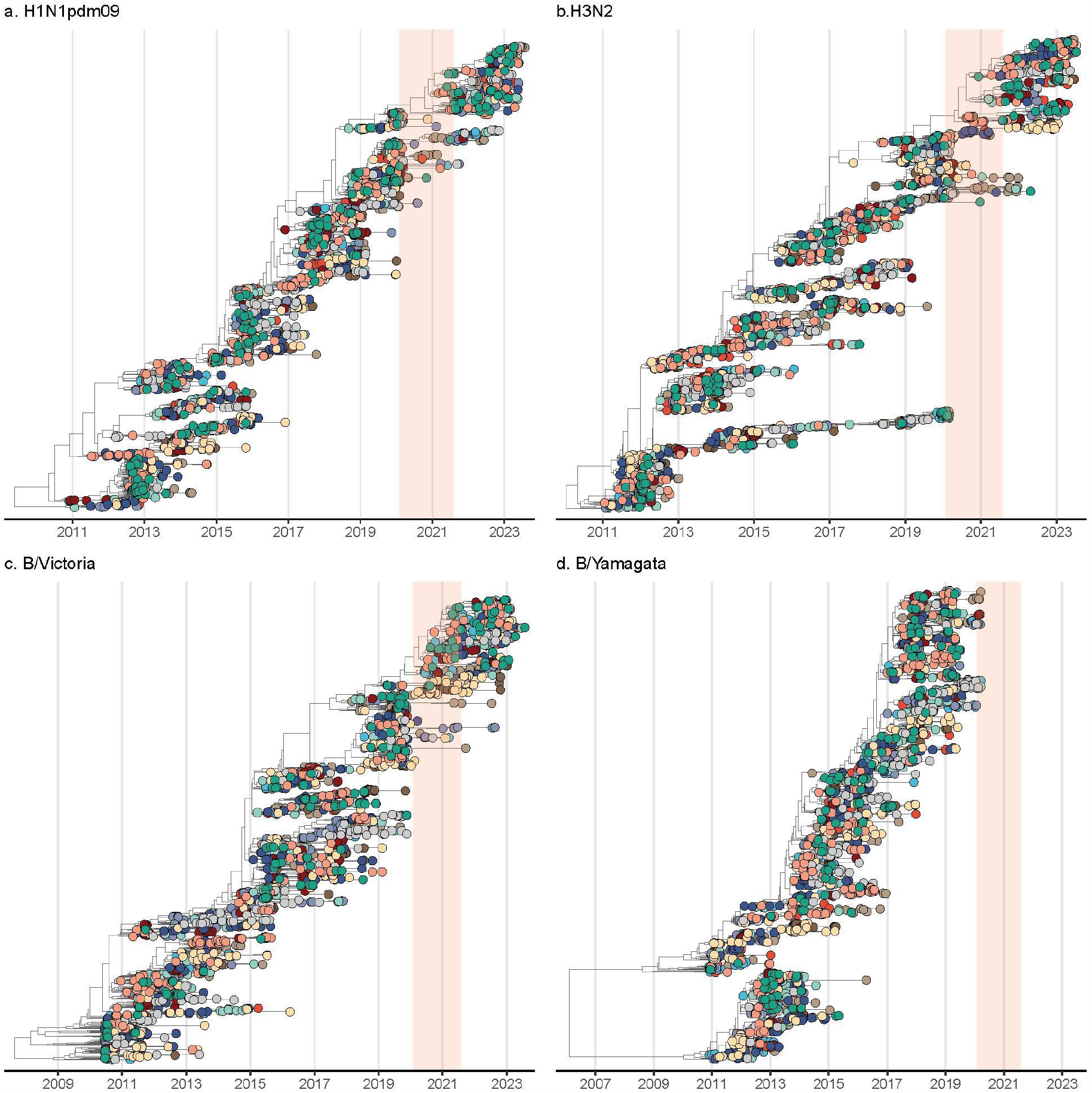
Maximum clade credibility (MCC) tree of seasonal influenza viruses dating back to 2011. Tips are annotated with the geographic regions whose colour legends are the same with Figure 3. The light orange shadow area represents the NPIs-intensive phase of COVID-19 pandemic, defined as spanning from 1 February 2020 to 31 July 2021.

**Extended Data Fig. 11.**
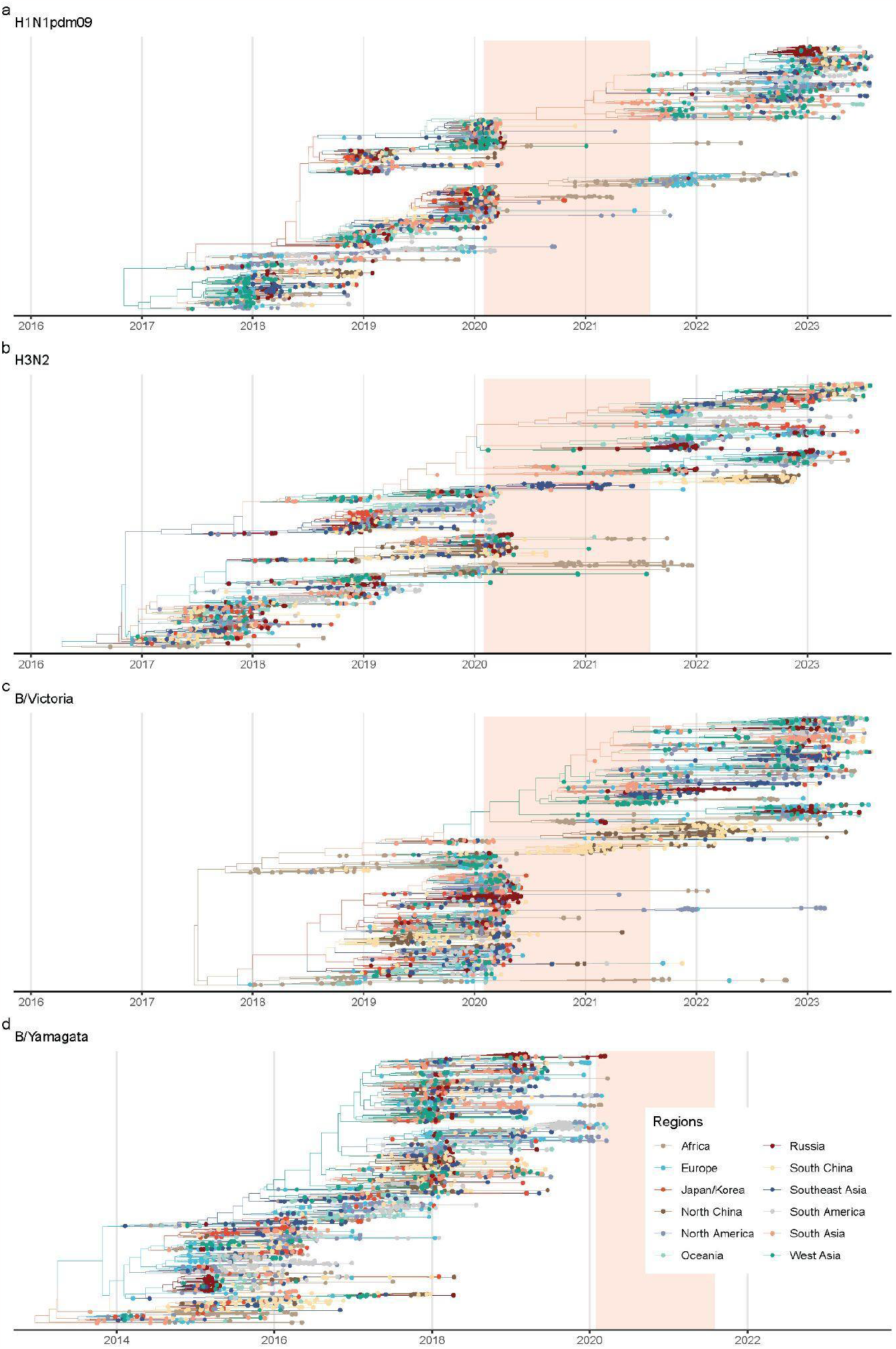
Maximum clade credibility (MCC) tree of the recently circulating clades of seasonal influenza viruses. The light orange shadow area represents the NPIs-intensive phase of COVID-19 pandemic, defined as spanning from 1 February 2020 to 31 July 2021.

**Extended Data Fig. 12.**
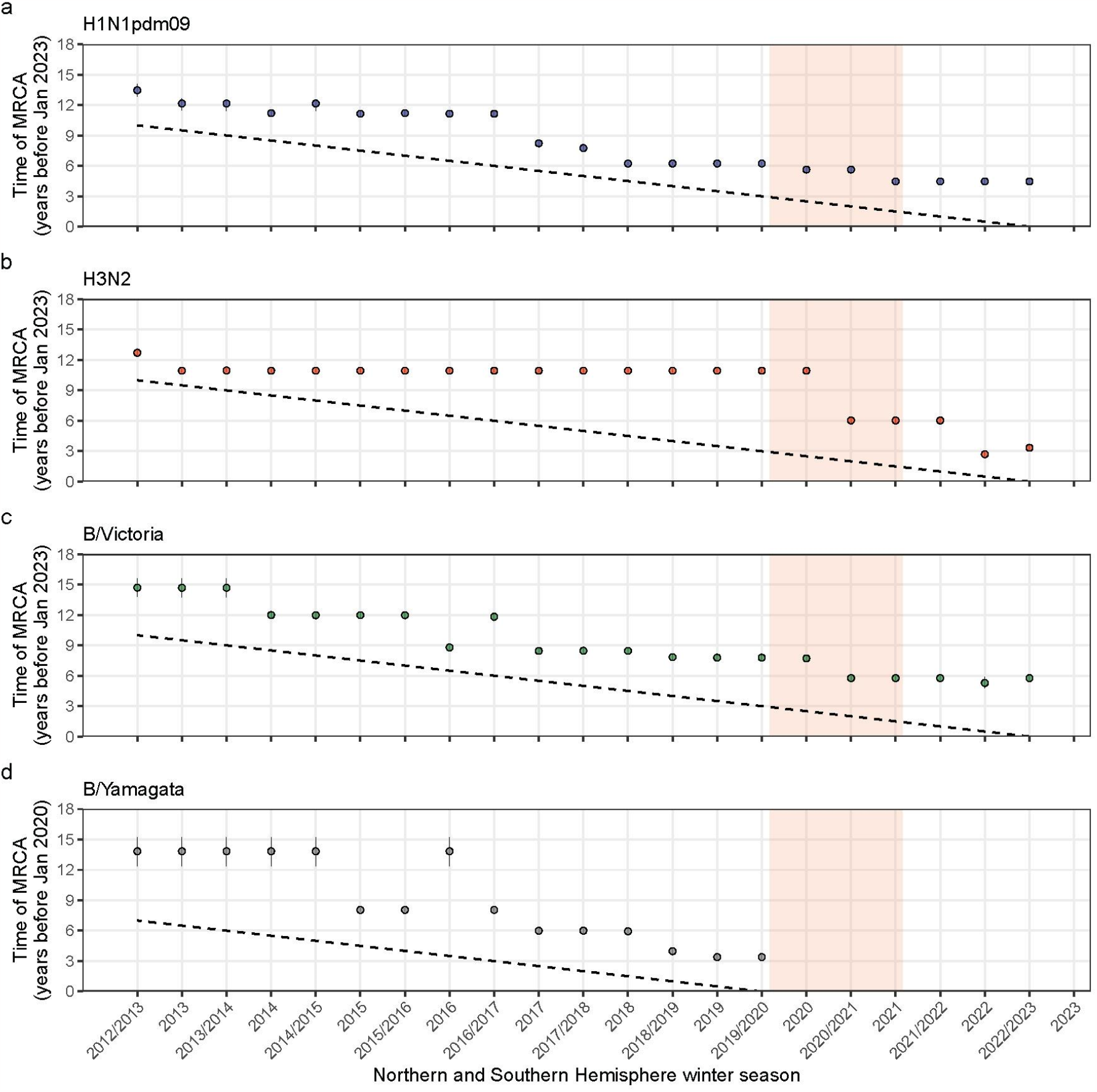
Lineage turnover of seasonal influenza viruses over influenza season. Lineage turnover was indicated by the time to the most recent common ancestor (TMRCA) of influenza viruses circulating during each season, in which mean values and 95% highest posterior density intervals were shown. The diagonal dotted line approximated the seasonal mid-point, which goes through 1st January of the Northern Hemisphere winter season and 1st July of the Southern Hemisphere winter season. Here, the Southern Hemisphere winter season was applied to Oceania, South America, and the Southern part of Africa.

## Supplementary Information

### Supplementary text: GLM diffusion predictors

Here, we included multiple categories of potential predictors of influenza spread in phylogeographic GLM framework and provided more details as follows:

#### Distance

We constructed a symmetric matrix of population-weighted distance among each pair of geographic regions. Within each geographic region we defined above, we first collected the population, centroid latitudes, and centroid longitudes at counties/states level, where we adopted state-level data as the spatial resolution for some countries with large land areas (Russia, Canada, China, United States, Brazil, Australia). The general formula for calculating the population-weighted distance between region *i* and region *j* is^68^:

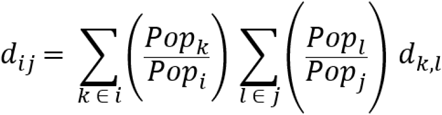

where Pop _k_ refers to the population of country k or state k belonging to region _i_, Pop _l_ the population of country _l_ or state _l_ belonging to region _j_, and d_k,l_ the distance between country k or state k and country l or state l.

#### Latitude

Here we calculated the region-level latitudes that are weighted by population at country/state level, in which the state-level spatial resolution was used in several countries with expansive land areas (Russia, Canada, China, United States, Brazil, and Australia).

#### Temperature/Precipitation/Relative humidity

We obtained meteorologic data from the ECMWF Reanalysis v5 (ERA5), which provides monthly average data for temperature, precipitation, relative humidity at a resolution of 0.25° × 0.25° from 1979 to present ^69^. To create interval-specific and region-specific ones, we averaged data from all grid cells within each pre-defined geographic region in each month and then averaged them during specific time intervals. Here, the first interval spanned from February 2018 to January 2020 (24 months), matching the duration of the third interval (from August 2021 to July 2023).

#### Air passenger traffic between and within regions

The origin-destination air traffic data up to July 2023 were provided by Official Airline Guide (OAG) Ltd. through a data sharing agreement (https://www.oag.com/). This dataset comprises 3,907 international airports and 1,469 domestic airports, with numbers of travel volume released on a monthly basis. Due to the delay of air traffic data release, the air traffic data in June and July 2023 are preliminarily adjusted while others are finally adjusted by OAG. We calculated the average monthly number of passengers on flights between each pair of regions per period we defined above. Additionally, we computed the monthly average number of air passengers travelled within each geographic region, which serves as a proxy for within-location air connectivity. The air traffic data used in the first, second, and third period ranged from February 2018 to January 2020 (24 months), from February 2020 to July 2021 (18 months), and from August 2021 to July 2023 (24 months), respectively.

#### COVID-19 stringency index

During the COVID-19 pandemic, the Oxford Coronavirus Government Response Tracker (OxCGRT) created a Stringency Index based on nine response indicators, spanning from 3 January 2020 to 31 December 2022^70^. We first calculated the mean COVID-19 stringency index for each country during each period. Due to data availability constraints, the first interval covered only 3 January 2020 to 31 January 2020; while the third interval ranged from 1 August 2021 to 31 December 2022. Subsequently, we computed a region-level index weighted by the population of each country.

#### Infectious disease vulnerability index

This index was developed to evaluate countries vulnerable to infectious disease outbreaks^71^. Here, we also computed a region-level index weighted by each country’s population.

#### Supplementary tables and figures

**Supplementary Fig. 1.**
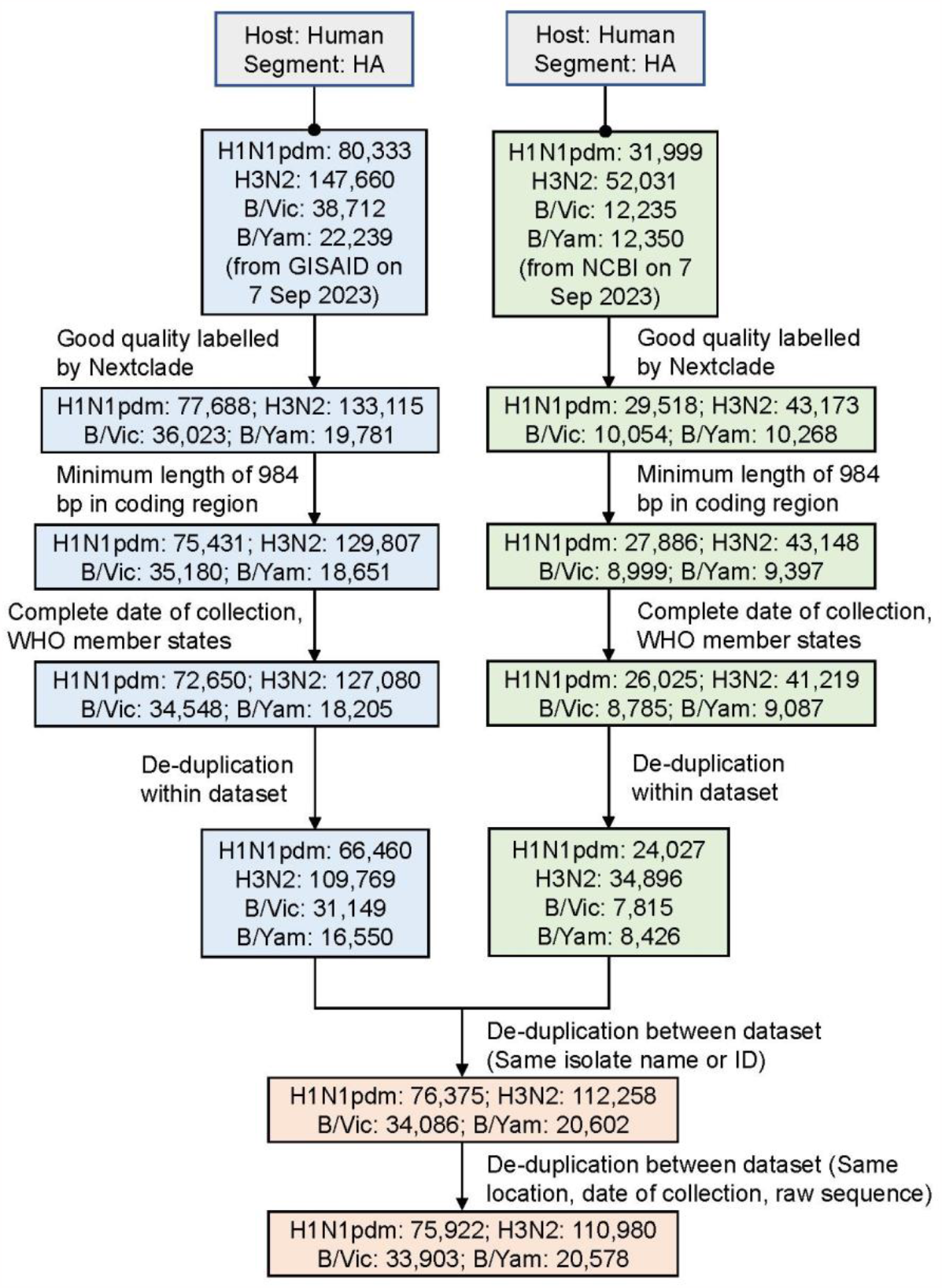
Flowchart of collating genetic data of seasonal influenza viruses.

**Supplementary Fig. 2.**
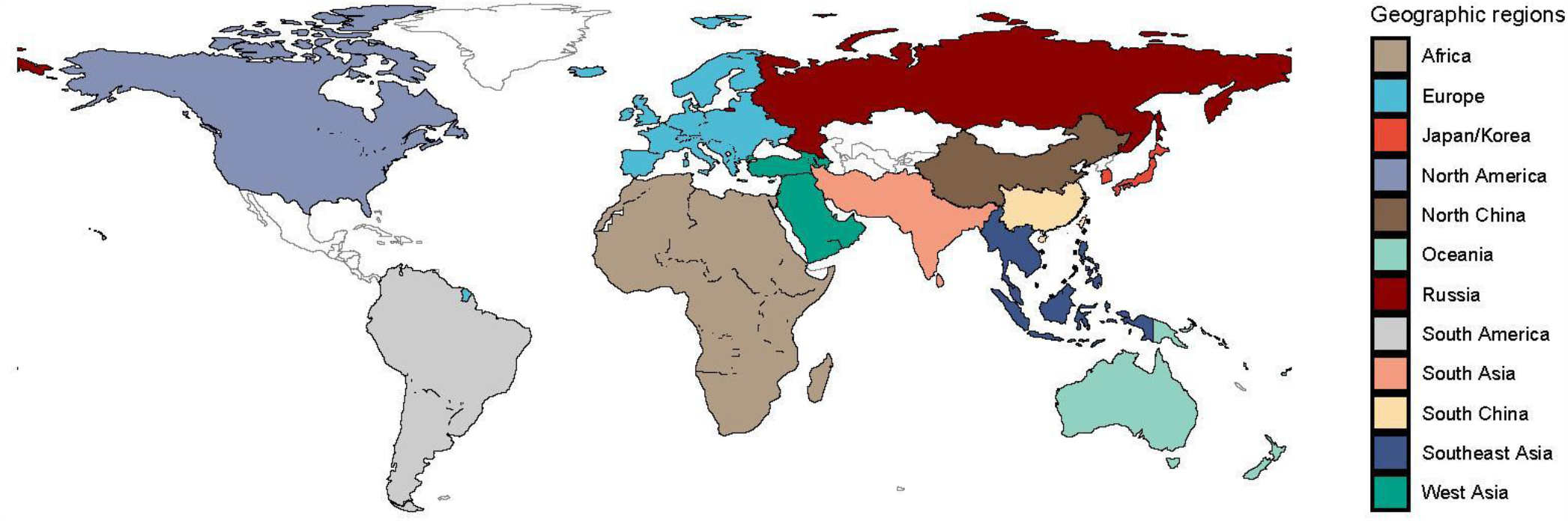
Geographic regions used in this study. Locations that are not included into this work are labelled as white.

**Supplementary Fig. 3.**
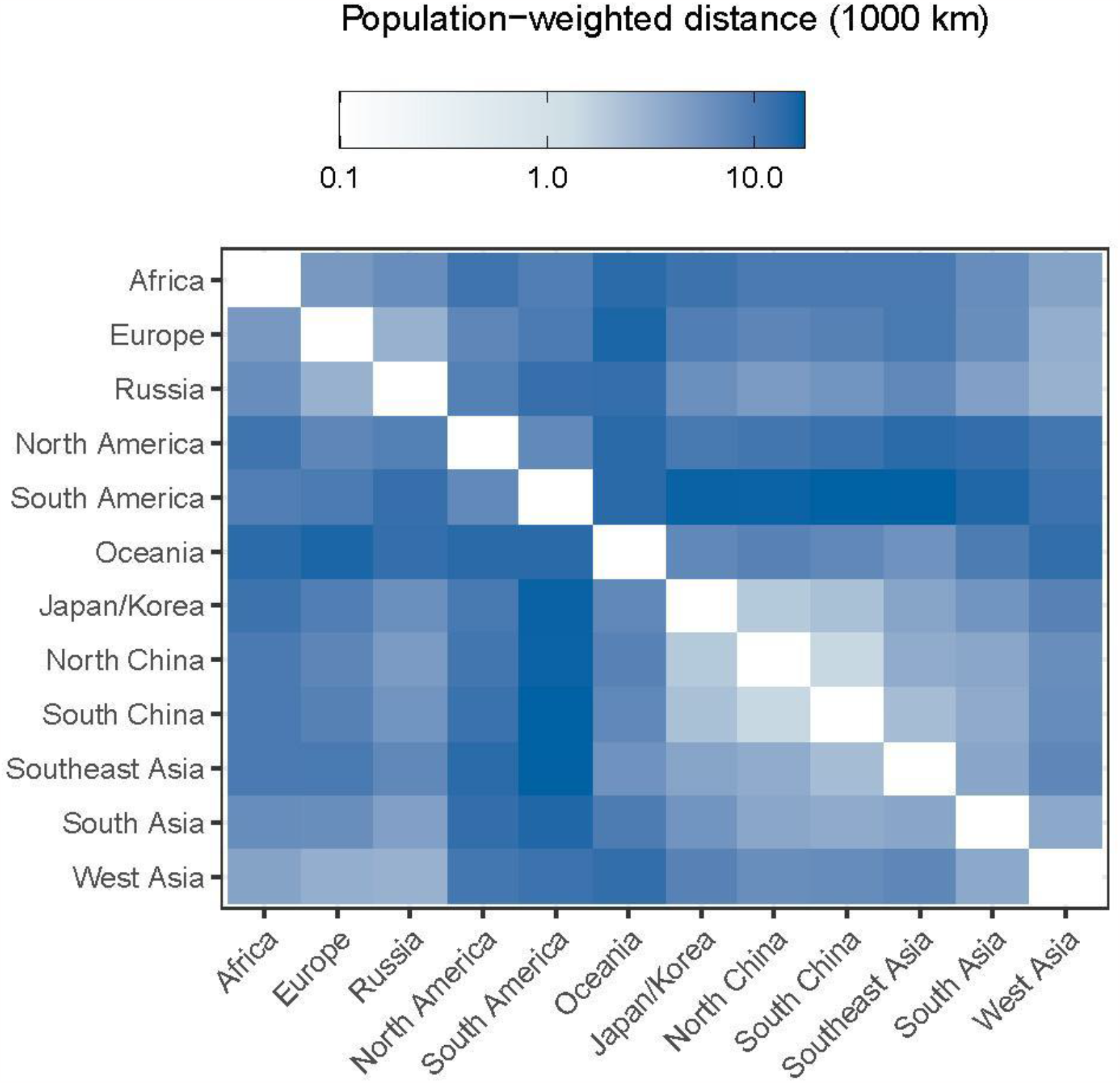
Population-weighted distance between geographic regions.

**Supplementary Fig. 4.**
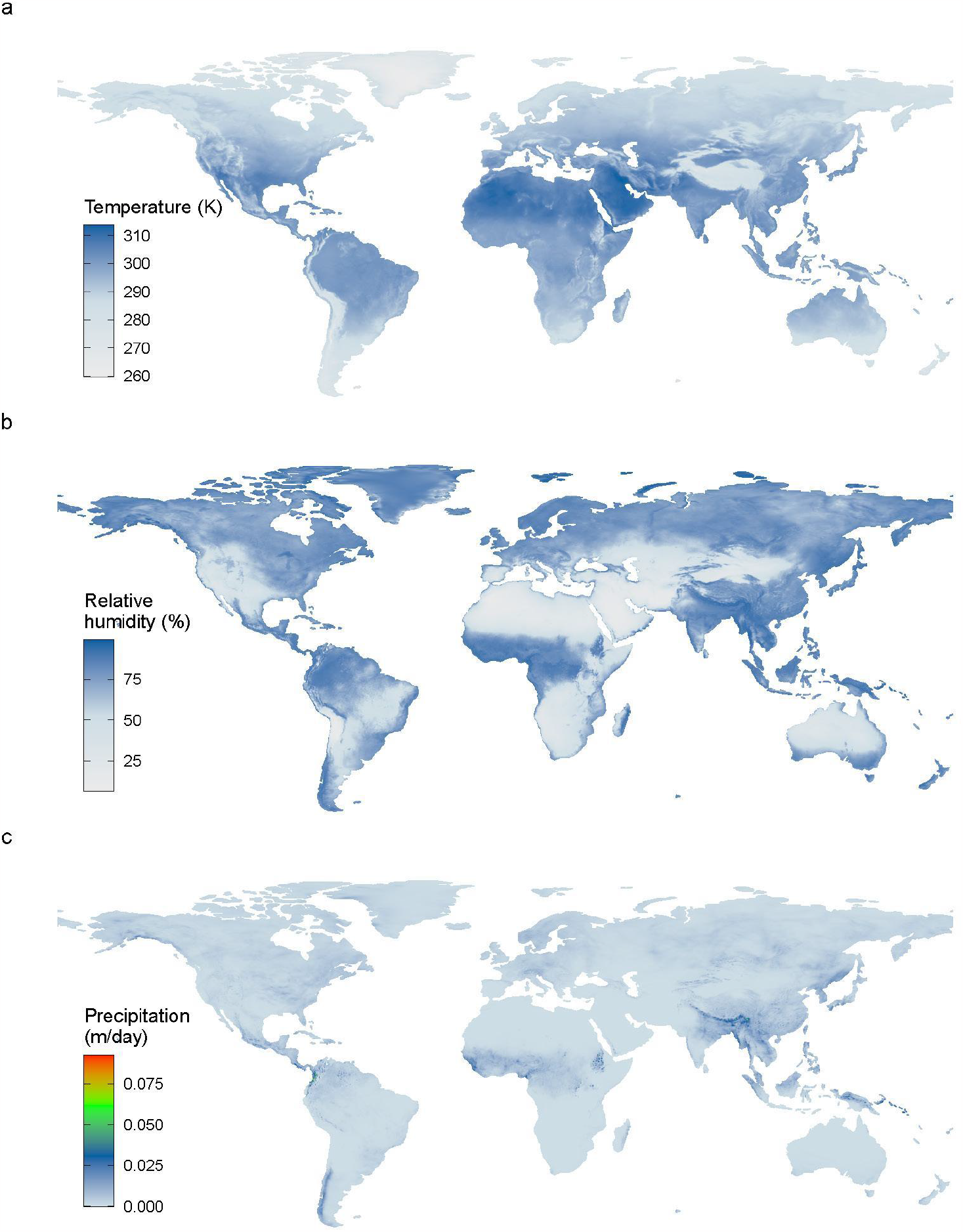
Mean temperature, relative humidity, precipitation at a spatial resolution of 0.25° × 0.25°. Here, we presented the data in August 2023 as an example.

**Supplementary Fig. 5.**
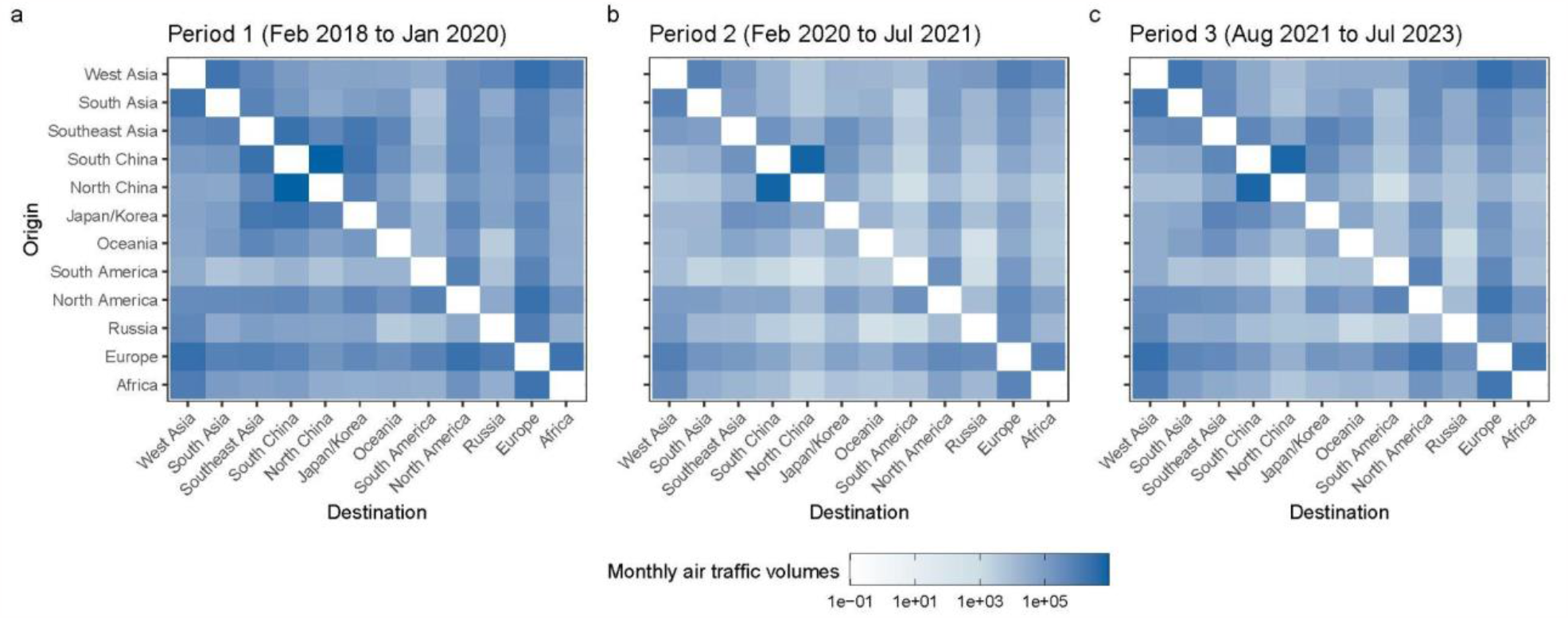
Monthly average of inter-regional air traffic matrices during the three periods.

**Supplementary Fig. 6.**
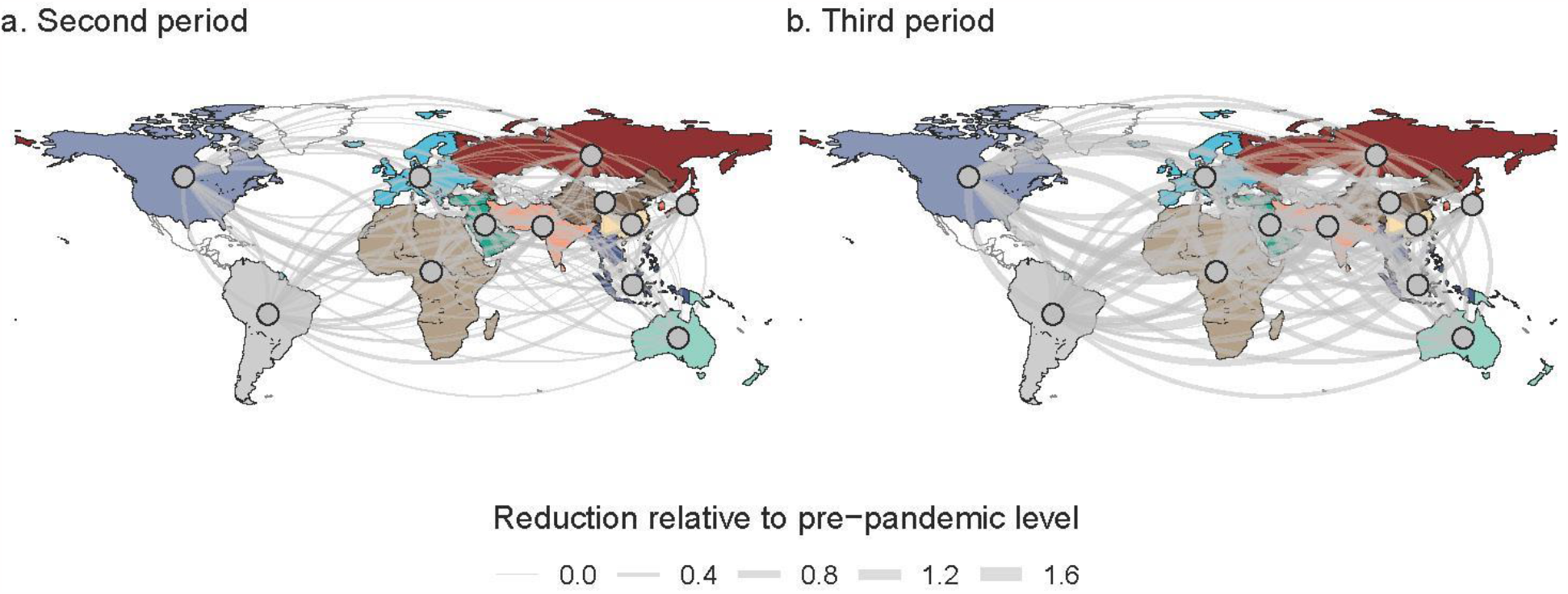
The air traffic reduction relative to pre-pandemic level in the second and third periods.

**Supplementary Fig. 7.**
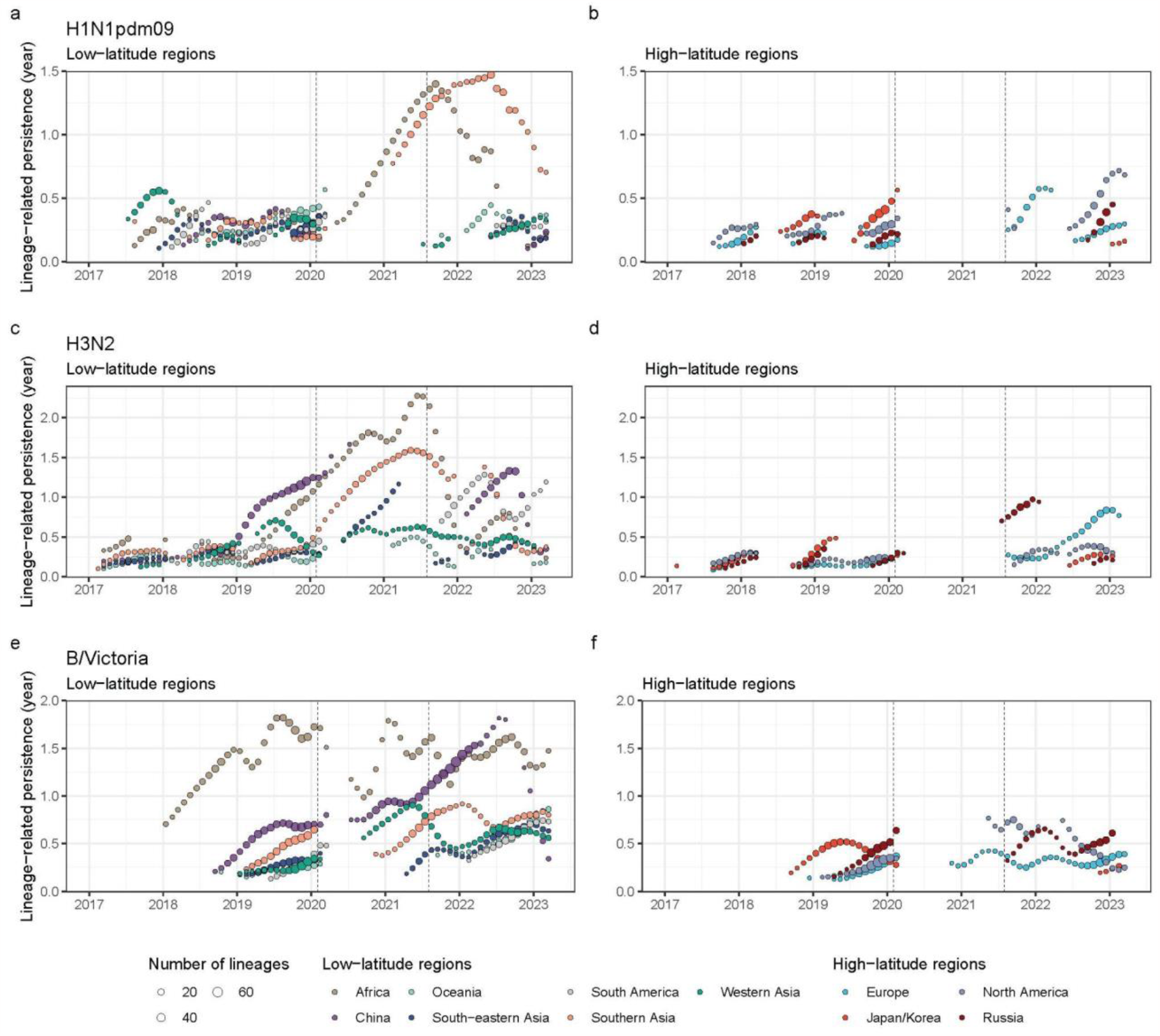
Lineage-associated persistence dynamic across geographic regions over time. This plot only showed the lineage-associated persistence at time points when the number of lineages circulating at that time points are over 5, in order to ensure the estimate of lineage-associated persistence will not be affected by the small size of lineages.

**Supplementary Fig. 8.**
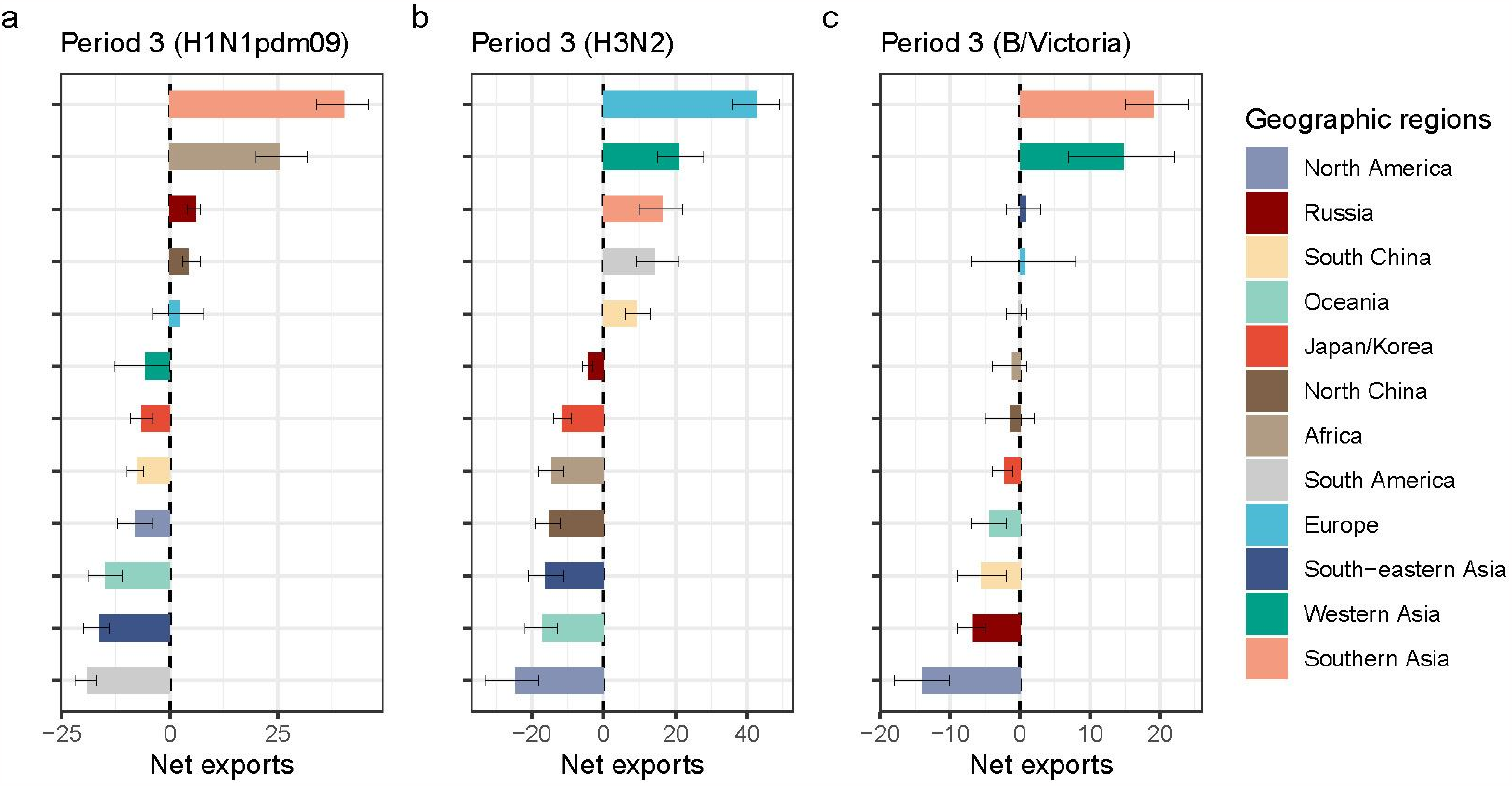
Net export dynamic of seasonal influenza in the third period under sub-sampling strategy accounting for the temporal variation.

**Supplementary Fig. 9.**
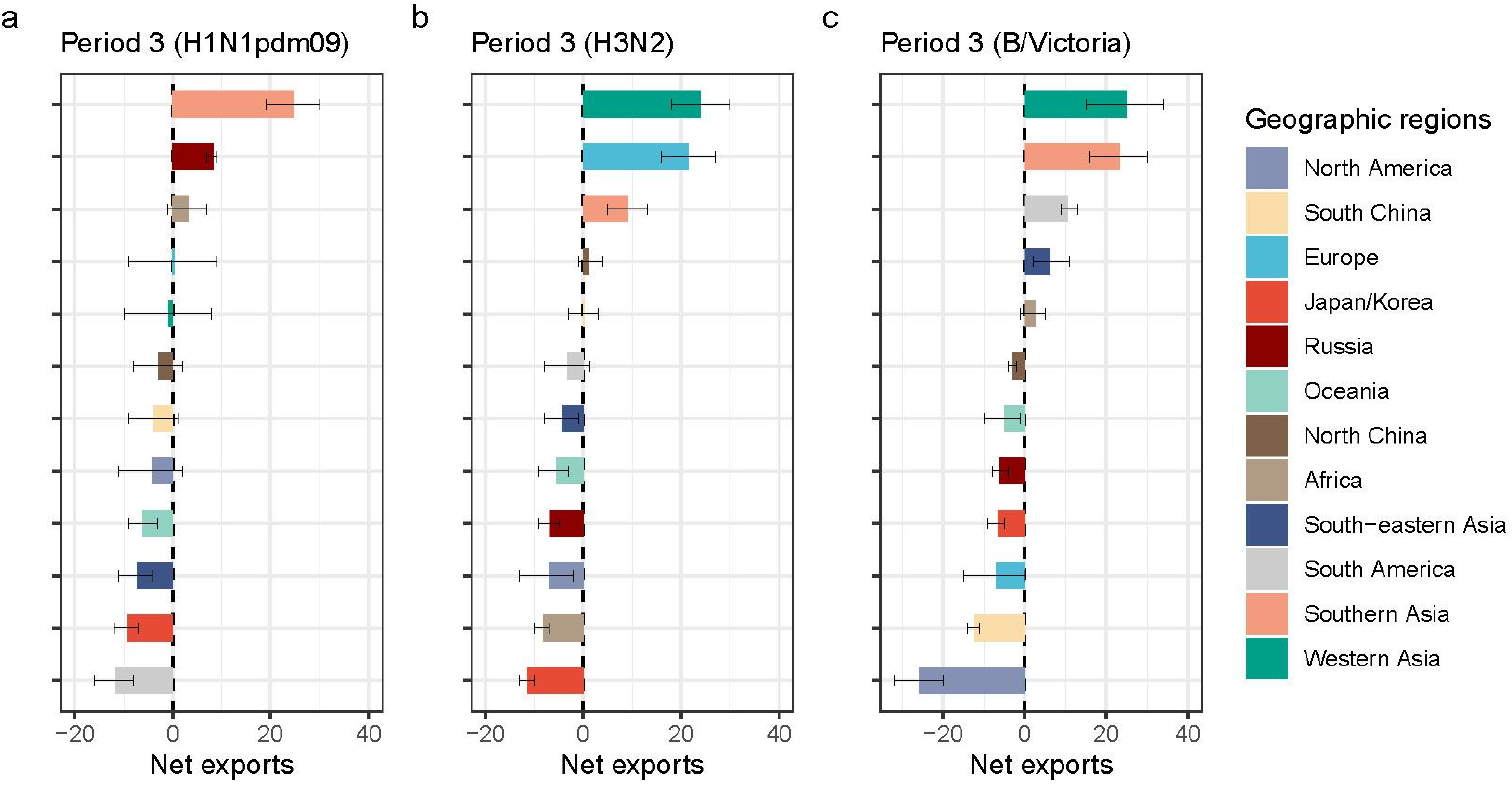
Net export dynamic of seasonal influenza in the third period when using region-specific cutoffs for defining the third period under the even sub-sampling strategy.

**Supplementary Fig. 10.**
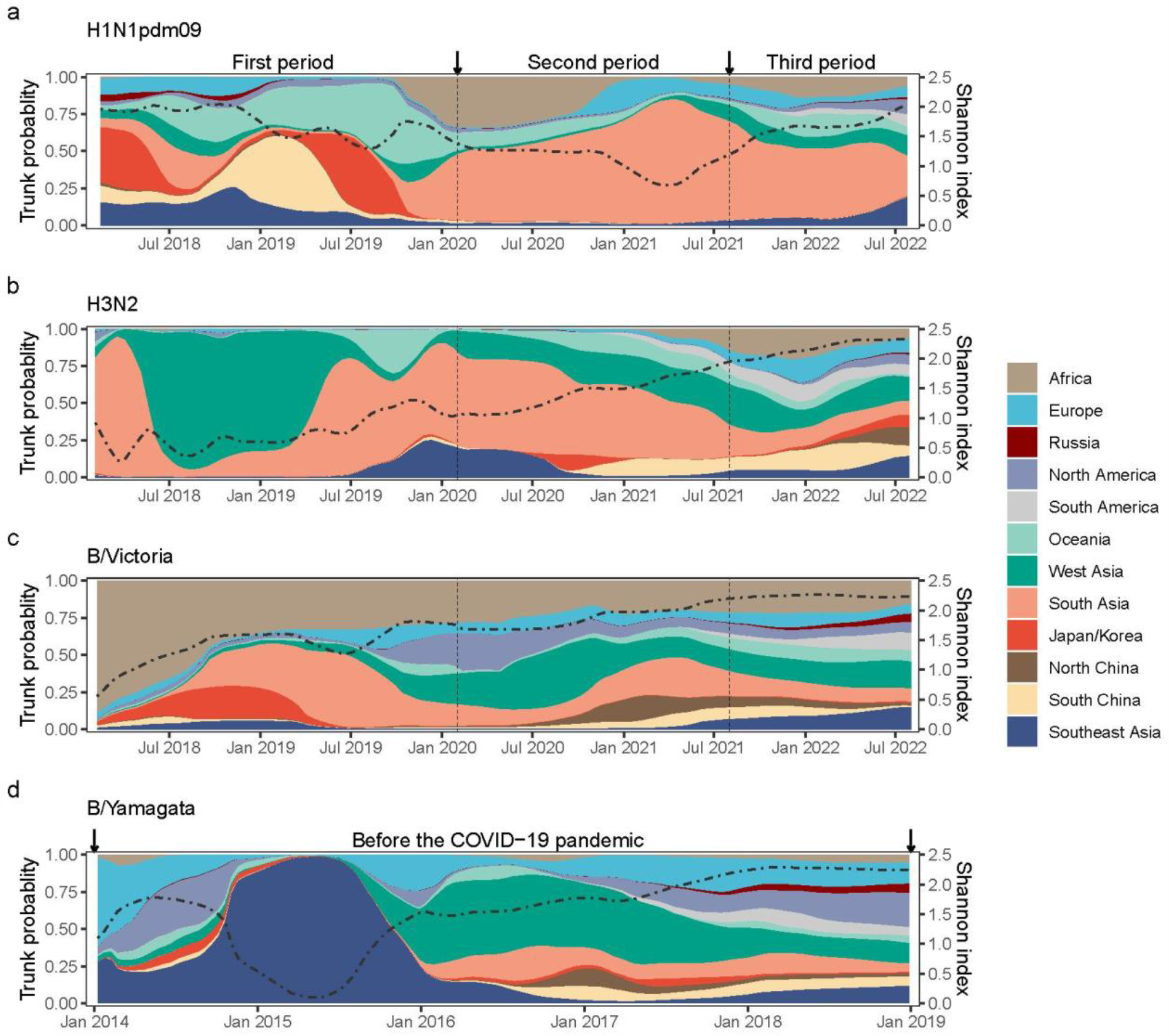
Inferred trunk location of phylogenetic trees under even sub-sampling strategy.

**Supplementary Fig. 11.**
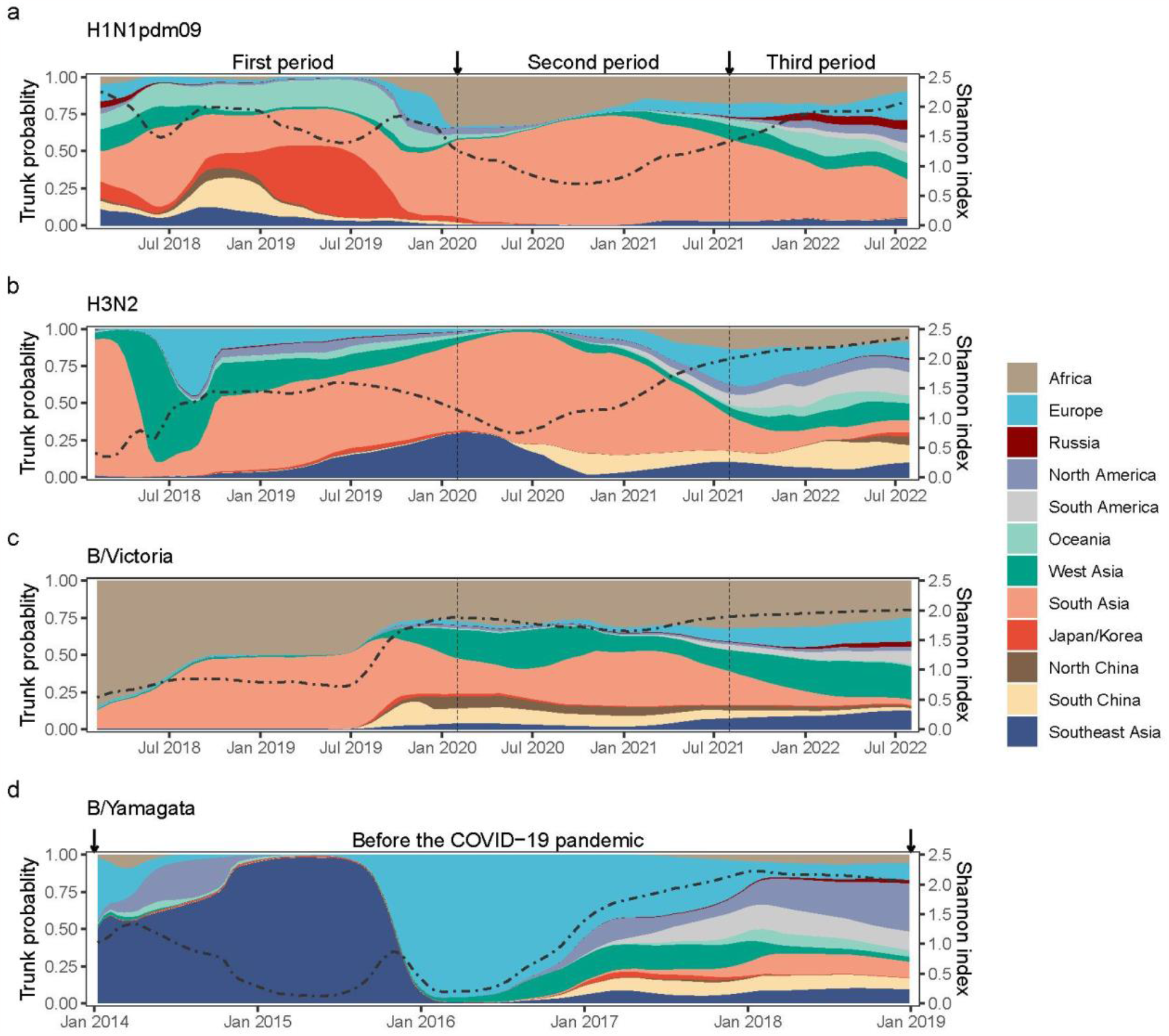
Inferred trunk location of phylogenetic trees under sub-sampling strategy accounting for the temporal variation.

**Supplementary Fig. 12.**
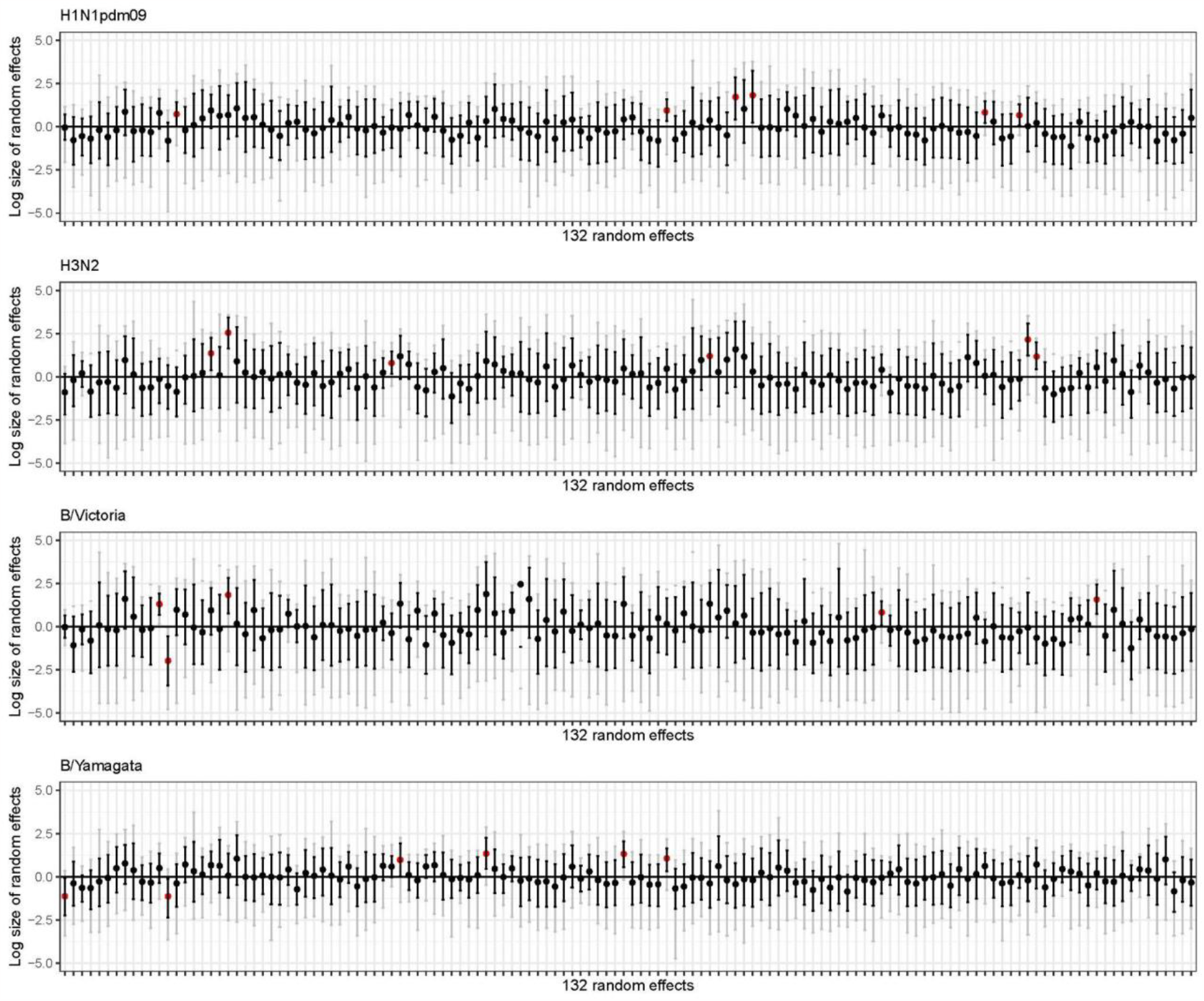
Posterior summary of the GLM random effects under even sampling strategy. Black points and error bars refer to the posterior mean and 95% HPD intervals, while grey error bars represent the minimum and maximum of the log size of random effects. Those random effects of transition rate with their 95% HPD intervals deviating 0 were marked as red points.

**Supplementary Fig. 13.**
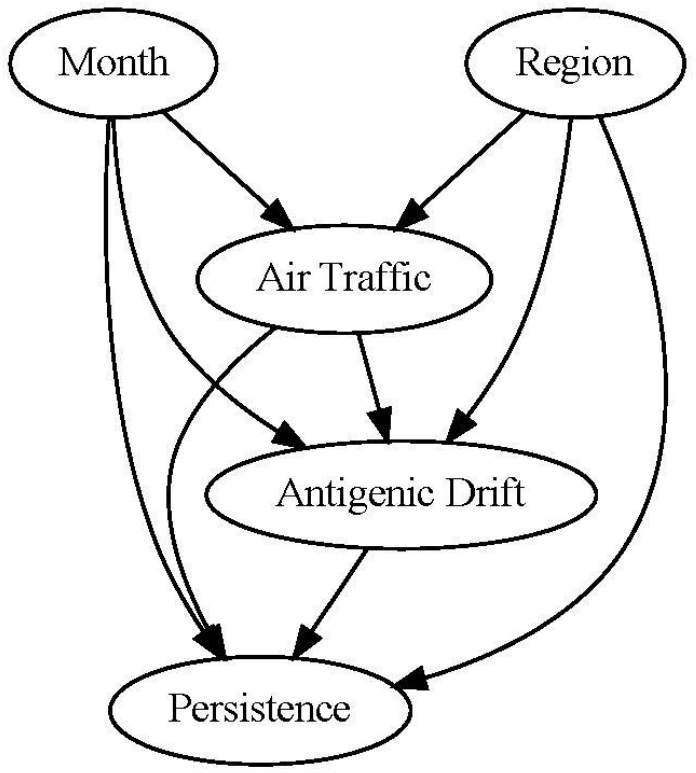
The directed acyclic graph (DAG) framework used in Bayesian hierarchical regression model.

**Supplementary Fig. 14.**
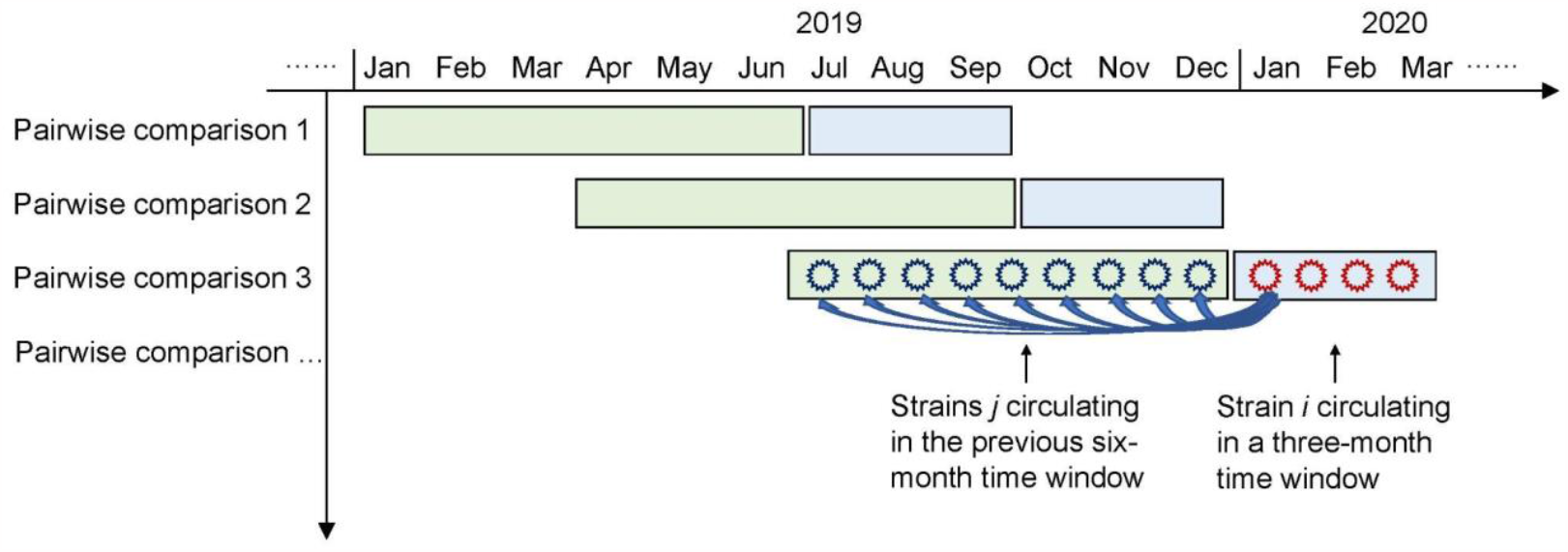
The conceptual representation of using genetic data to measure cross-immunity to indicate the extent of antigenic drift.

**Supplementary Table 1.**
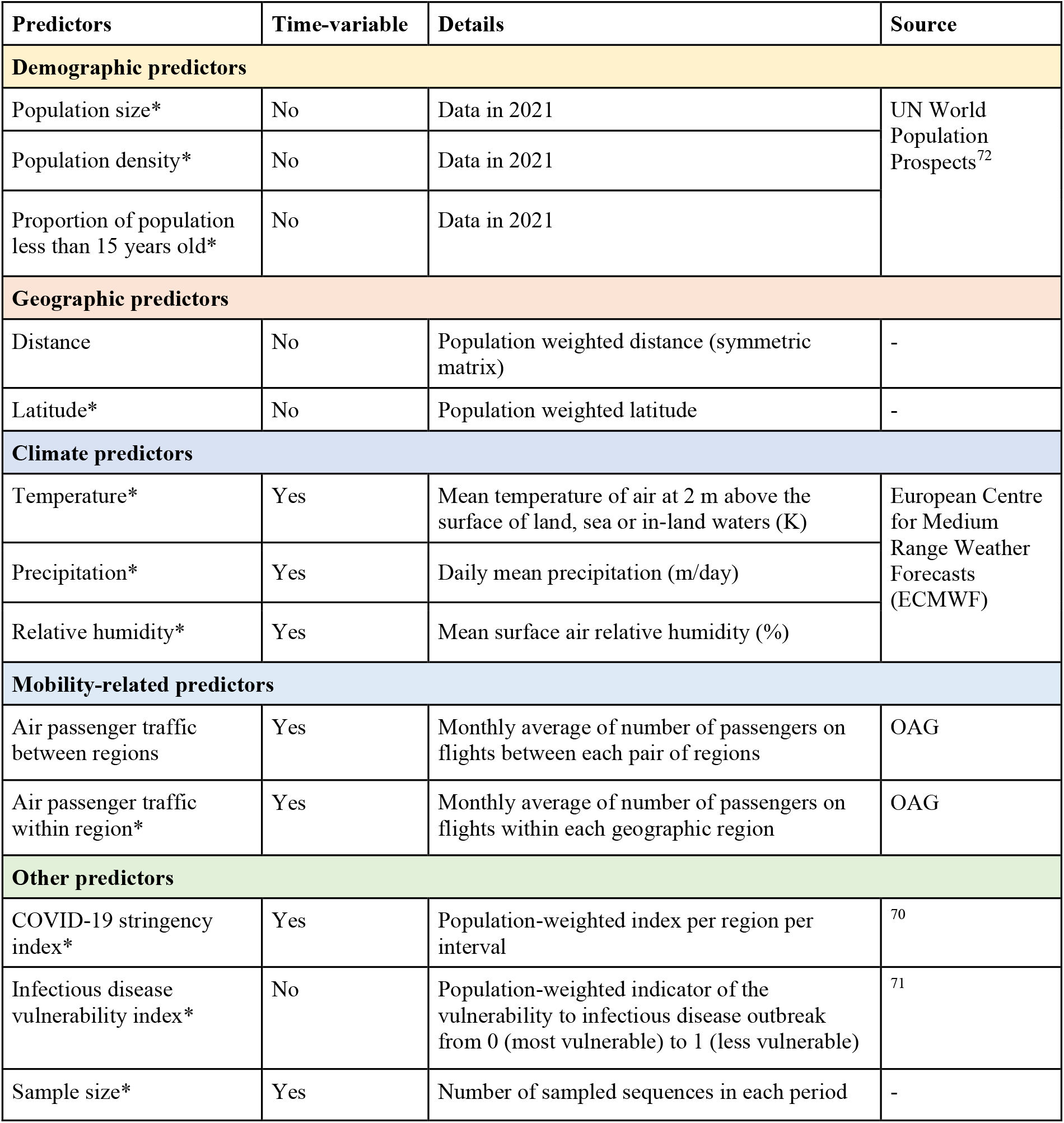
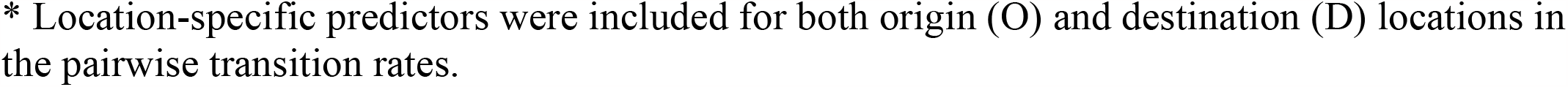
Potential predictors of spatial spread of seasonal influenza included in this study.

| Predictors | Time-variable | Details | Source |
| --- | --- | --- | --- |
| Demographic predictors |  |  |  |
| Population size* | No | Data in 2021 | UN World Population Prospects<sup>72</sup> |
| Population density* | No | Data in 2021 |  |
| Proportion of population less than 15 years old* | No | Data in 2021 |  |
| Geographic predictors |  |  |  |
| Distance | No | Population weighted distance (symmetric matrix) | - |
| Latitude* | No | Population weighted latitude | - |
| Climate predictors |  |  |  |
| Temperature* | Yes | Mean temperature of air at 2 m above the surface of land, sea or in-land waters (K) | European Centre for Medium Range Weather Forecasts (ECMWF) |
| Precipitation* | Yes | Daily mean precipitation (m/day) |  |
| Relative humidity* | Yes | Mean surface air relative humidity (%) |  |
| Mobility-related predictors |  |  |  |
| Air passenger traffic between regions | Yes | Monthly average of number of passengers on flights between each pair of regions | OAG |
| Air passenger traffic within region* | Yes | Monthly average of number of passengers on flights within each geographic region | OAG |
| Other predictors |  |  |  |
| COVID-19 stringency index* | Yes | Population-weighted index per region per interval | 70 |
| Infectious disease vulnerability index* | No | Population-weighted indicator of the vulnerability to infectious disease outbreak from 0 (most vulnerable) to 1 (less vulnerable) | 71 |
| Sample size* | Yes | Number of sampled sequences in each period | - |

\* Location-specific predictors were included for both origin (O) and destination (D) locations in the pairwise transition rates.

**Supplementary Table 2.** Predictors of spatial spread of seasonal influenza used in GLM framework in this study.

| **Predictors** | **Africa** | **Europe** | **Russia** | **North America** | **South America** | **Oceania** | **Japan/Korea** | **North China** | **South China** | **Southeast Asia** | **South Asia** | **West Asia** |
| --- | --- | --- | --- | --- | --- | --- | --- | --- | --- | --- | --- | --- |
| Pop size (million) | 1391.8 | 598.0 | 145.1 | 375.2 | 434.0 | 43.6 | 176.4 | 575.9 | 865.9 | 675.8 | 1989.5 | 284.6 |
| Pop density (pop per km<sup>2</sup>) | 46.9 | 103.9 | 8.9 | 20.7 | 25.0 | 5.1 | 381.8 | 84.1 | 331.1 | 153.3 | 311.2 | 59.5 |
| Prop of pop under 15 (%) | 40.4 | 15.4 | 17.7 | 18.0 | 22.5 | 23.1 | 11.8 | 17.8 | 17.9 | 24.8 | 27.4 | 28.8 |
| Latitude | 5.3 | 48.5 | 54.3 | 39.1 | -14.9 | -27.3 | 36.1 | 38.1 | 27.9 | 8.0 | 22.7 | 31.6 |
| Temperature 1 (K)* | 297.9 | 281.7 | 269.4 | 272.9 | 294.7 | 295.4 | 285.8 | 277.3 | 289.3 | 298.6 | 293.9 | 295.7 |
| Temperature 2 (K) | 298.2 | 282.0 | 270.8 | 273.7 | 294.4 | 294.5 | 285.9 | 278.5 | 289.9 | 298.7 | 294.5 | 296.2 |
| Temperature 3 (K) | 297.9 | 281.7 | 269.3 | 273.3 | 294.5 | 294.6 | 285.9 | 277.5 | 289.5 | 298.5 | 294.0 | 295.5 |
| Precipitation 1 (mm/day) | 1.9 | 2.3 | 1.5 | 2.0 | 4.6 | 1.5 | 4.6 | 1.4 | 4.1 | 6.8 | 2.5 | 0.8 |
| Precipitation 2 (mm/day) | 1.7 | 2.5 | 1.5 | 1.9 | 4.4 | 2.0 | 4.9 | 1.5 | 4.6 | 7.4 | 2.5 | 0.7 |
| Precipitation 3 (mm/day) | 1.7 | 2.4 | 1.5 | 1.9 | 4.5 | 2.0 | 4.7 | 1.4 | 4.0 | 7.6 | 2.4 | 0.7 |
| Relative humidity 1 (%) | 51.1 | 76.4 | 75.0 | 72.7 | 73.6 | 48.4 | 74.1 | 54.7 | 77.0 | 81.5 | 57.1 | 46.3 |
| Relative humidity 2 (%) | 49.9 | 75.3 | 73.3 | 70.8 | 72.8 | 53.8 | 74.5 | 53.2 | 76.6 | 81.6 | 55.8 | 42.6 |
| Relative humidity 3 (%) | 50.0 | 76.4 | 74.8 | 71.6 | 72.5 | 55.0 | 74.7 | 54.2 | 75.3 | 82.5 | 57.4 | 44.4 |
| Air traffic within region 1 (million) | 4.2 | 51.5 | 3.9 | 54.1 | 13.5 | 6.3 | 10.0 | 6.8 | 19.0 | 21.8 | 11.3 | 8.0 |
| Air traffic within region 2 (million) | 1.8 | 13.7 | 3.7 | 25.7 | 5.3 | 2.6 | 5.7 | 5.1 | 14.7 | 8.8 | 5.7 | 3.3 |
| Air traffic within region 3 (million) | 3.8 | 41.4 | 4.8 | 50.0 | 12.1 | 4.5 | 8.5 | 3.9 | 12.5 | 13.6 | 10.9 | 6.5 |
| Stringency index 1 | 0.7 | 1.1 | 0.6 | 0.1 | 1.1 | 1.8 | 1.7 | 23.1 | 23.1 | 4.5 | 2.1 | 0.5 |
| Stringency index 2 | 51.1 | 60.4 | 47.9 | 61.9 | 67.6 | 53.8 | 44.7 | 71.9 | 70.7 | 64.2 | 68.8 | 59.3 |
| Stringency index 3 | 30.5 | 26.6 | 36.6 | 37.4 | 34.0 | 38.3 | 38.3 | 77.9 | 76.3 | 46.1 | 45.3 | 30.4 |
| Infectious disease vulnerability index | 0.34 | 0.83 | 0.64 | 0.93 | 0.67 | 0.76 | 0.91 | 0.66 | 0.67 | 0.58 | 0.46 | 0.58 |

\* The number after the predictors represents the three time intervals defined in this study.

**Supplementary Table 3.** The sample size of each influenza virus per period per geographic region per sub-sampling strategy.

| Sub-sampling strategy* | Periods | Africa | Europe | Russia | North America | South America | Oceania | Japan/Korea | North China | South China | Southeast Asia | South Asia | West Asia |
| --- | --- | --- | --- | --- | --- | --- | --- | --- | --- | --- | --- | --- | --- |
| **H1N1pdm09** |  |  |  |  |  |  |  |  |  |  |  |  |  |
| Even | Period 1 | 104 | 116 | 81 | 121 | 72 | 83 | 92 | 66 | 84 | 91 | 90 | 123 |
| Even | Period 2 | 56 | 21 | 20 | 19 | 31 | 19 | 8 | 15 | 17 | 11 | 21 | 10 |
| Even | Period 3 | 72 | 78 | 53 | 70 | 51 | 54 | 21 | 18 | 21 | 51 | 72 | 74 |
| Temporal | Period 1 | 94 | 98 | 80 | 119 | 102 | 91 | 65 | 39 | 73 | 102 | 92 | 102 |
| Temporal | Period 2 | 97 | 26 | 24 | 35 | 30 | 28 | 24 | 11 | 13 | 25 | 42 | 23 |
| Temporal | Period 3 | 75 | 89 | 30 | 59 | 48 | 66 | 14 | 13 | 18 | 38 | 66 | 49 |
| Spatiotemporal | Period 1 | 30 | 237 | 44 | 453 | 39 | 35 | 18 | 18 | 56 | 24 | 35 | 69 |
| Spatiotemporal | Period 2 | 87 | 49 | 17 | 197 | 2 | 2 | 3 | - | 2 | 1 | 11 | 6 |
| Spatiotemporal | Period 3 | 40 | 182 | 18 | 120 | 54 | 33 | 1 | 28 | 53 | 5 | 32 | 10 |
| **H3N2** |  |  |  |  |  |  |  |  |  |  |  |  |  |
| Even | Period 1 | 84 | 99 | 72 | 105 | 65 | 95 | 98 | 76 | 94 | 86 | 82 | 76 |
| Even | Period 2 | 42 | 12 | 18 | 7 | 21 | 19 | 15 | 10 | 19 | 43 | 36 | 20 |
| Even | Period 3 | 59 | 70 | 69 | 70 | 69 | 64 | 46 | 42 | 48 | 54 | 61 | 68 |
| Temporal | Period 1 | 57 | 98 | 39 | 111 | 52 | 91 | 56 | 17 | 83 | 117 | 61 | 71 |
| Temporal | Period 2 | 48 | 14 | 8 | 15 | 2 | 7 | 1 | - | 5 | 48 | 43 | 5 |
| Temporal | Period 3 | 97 | 113 | 44 | 111 | 105 | 89 | 51 | 37 | 78 | 103 | 69 | 88 |
| Spatiotemporal | Period 1 | 36 | 145 | 22 | 278 | 24 | 87 | 22 | 19 | 86 | 53 | 43 | 38 |
| Spatiotemporal | Period 2 | 45 | 23 | 6 | 17 | - | 5 | - | - | 6 | 51 | 40 | 7 |
| Spatiotemporal | Period 3 | 23 | 231 | 17 | 492 | 52 | 22 | 13 | 16 | 53 | 21 | 25 | 15 |
| **B/Victoria** |  |  |  |  |  |  |  |  |  |  |  |  |  |
| Even | Period 1 | 104 | 59 | 52 | 65 | 55 | 62 | 59 | 49 | 81 | 67 | 71 | 66 |
| Even | Period 2 | 38 | 42 | 37 | 28 | 26 | 16 | 21 | 33 | 66 | 34 | 28 | 37 |
| Even | Period 3 | 92 | 90 | 69 | 90 | 62 | 68 | 22 | 59 | 78 | 81 | 63 | 80 |
| Temporal | Period 1 | 140 | 94 | 76 | 119 | 57 | 80 | 87 | 34 | 106 | 106 | 102 | 106 |
| Temporal | Period 2 | 56 | 42 | 36 | 36 | 34 | 13 | 32 | 21 | 58 | 19 | 33 | 31 |
| Temporal | Period 3 | 70 | 61 | 34 | 36 | 46 | 30 | 9 | 21 | 54 | 42 | 33 | 40 |
| Spatiotemporal | Period 1 | 79 | 73 | 17 | 592 | 8 | 44 | 26 | 24 | 166 | 35 | 30 | 15 |
| Spatiotemporal | Period 2 | 35 | 88 | 17 | 151 | 5 | - | 4 | 12 | 75 | 6 | 11 | 7 |
| Spatiotemporal | Period 3 | 38 | 131 | 12 | 37 | 66 | 21 | - | 34 | 69 | 24 | 25 | 17 |
| **B/Yamagata** |  |  |  |  |  |  |  |  |  |  |  |  |  |
| Even | - | 180 | 221 | 114 | 210 | 191 | 180 | 167 | 96 | 173 | 190 | 166 | 141 |
| Temporal | - | 143 | 218 | 80 | 338 | 240 | 153 | 158 | 63 | 170 | 218 | 92 | 130 |
| Spatiotemporal | - | 52 | 450 | 22 | 890 | 118 | 97 | 63 | 27 | 140 | 65 | 38 | 42 |

\* “Even” denotes even sampling strategy; “Temporal” denotes sampling strategy accounting for the temporal variation; “Spatiotemporal” denotes sampling strategy accounting for the temporal and spatial variation.

**Supplementary Table 4.**
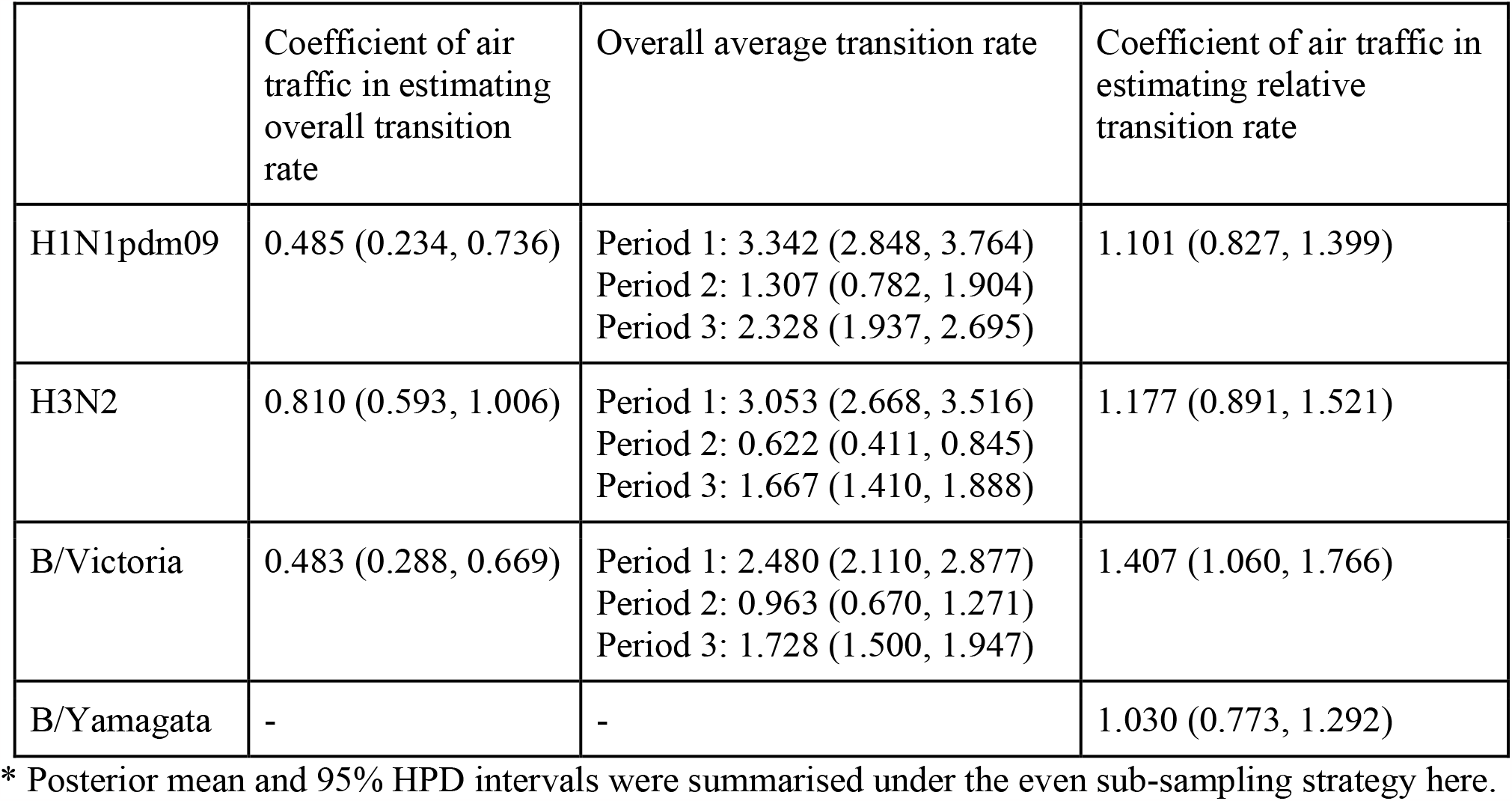
Parameter estimates under the 3-period phylogeographic model with air traffic data as the sole predictor of relative and overall transition rates

|  | Coefficient of air traffic in estimating overall transition rate | Overall average transition rate | Coefficient of air traffic in estimating relative transition rate |
| --- | --- | --- | --- |
| H1N1pdm09 | 0.485 (0.234, 0.736) | Period 1: 3.342 (2.848, 3.764)<br>Period 2: 1.307 (0.782, 1.904)<br>Period 3: 2.328 (1.937, 2.695) | 1.101 (0.827, 1.399) |
| H3N2 | 0.810 (0.593, 1.006) | Period 1: 3.053 (2.668, 3.516)<br>Period 2: 0.622 (0.411, 0.845)<br>Period 3: 1.667 (1.410, 1.888) | 1.177 (0.891, 1.521) |
| B/Victoria | 0.483 (0.288, 0.669) | Period 1: 2.480 (2.110, 2.877)<br>Period 2: 0.963 (0.670, 1.271)<br>Period 3: 1.728 (1.500, 1.947) | 1.407 (1.060, 1.766) |
| B/Yamagata | - | - | 1.030 (0.773, 1.292) |

\* Posterior mean and 95% HPD intervals were summarised under the even sub-sampling strategy here.

**Supplementary Table 5.**
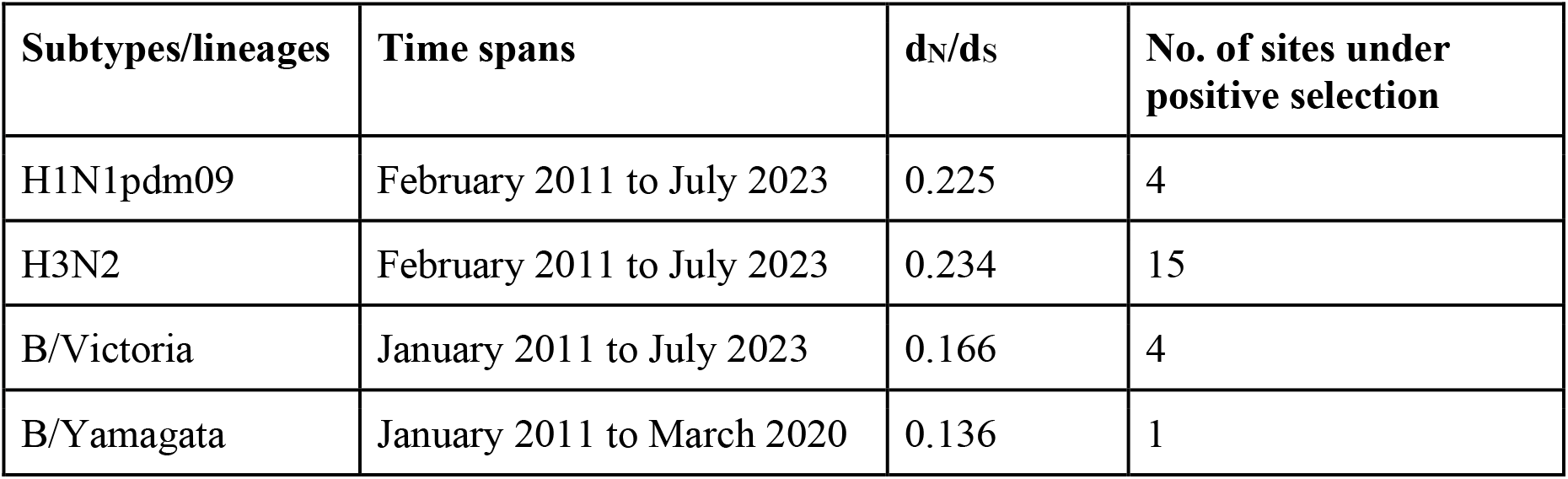
Selection pressures (d_N_/d_S_ ratio) and number of sites under positive selection of influenza viruses.

| Subtypes/lineages | Time spans | d<sub>N</sub>/d<sub>S</sub> | No. of sites under positive selection |
| --- | --- | --- | --- |
| H1N1pdm09 | February 2011 to July 2023 | 0.225 | 4 |
| H3N2 | February 2011 to July 2023 | 0.234 | 15 |
| B/Victoria | January 2011 to July 2023 | 0.166 | 4 |
| B/Yamagata | January 2011 to March 2020 | 0.136 | 1 |

